# The UK Biobank Mental Health Enhancement 2022: Methods and Results

**DOI:** 10.1101/2024.11.21.24317700

**Authors:** Katrina A.S. Davis, Jonathan R.I. Coleman, Mark Adams, Gerome Breen, Na Cai, Helena Davies, Kelly Davies, Alexandru Dregan, Thalia C. Eley, Elaine Fox, Jo Holliday, Christopher Huebel, Ann John, Aliyah S. Kassam, Matthew J. Kempton, William Lee, Danyang Li, Jared Maina, Rose McCabe, Andrew M. McIntosh, Sian Oram, Marcus Richards, Megan Skelton, Fenella Starkey, Abigail R ter Kuile, Laura M Thronton, Rujia Wang, Zhaoying Yu, Johan Zvrskovec, Matthew Hotopf

## Abstract

**Background:** This paper introduces the UK Biobank (UKB) second mental health questionnaire (MHQ2), describes its design, the respondents and some notable findings. UKB is a large cohort study with over 500,000 volunteer participants aged 40-69 years when recruited in 2006-2010. It is an important resource of extensive health, genetic and biomarker data. Enhancements to UKB enrich the data available. MHQ2 is an enhancement designed to enable and facilitate research with psychosocial and mental health aspects.

**Methods:** UKB sent participants a link to MHQ2 by email in October-November 2022. The MHQ2 was designed by a multi-institutional consortium to build on MHQ1. It characterises lifetime depression further, adds data on panic disorder and eating disorders, repeats ‘current’ mental health measures and updates information about social circumstances. It includes established measures, such as the PHQ-9 for current depression and CIDI-SF for lifetime panic, as well as bespoke questions. Algorithms and R code were developed to facilitate analysis.

**Results:** At the time of analysis, MHQ2 results were available for 169,253 UKB participants, of whom 111,275 had also completed the earlier MHQ1. Characteristics of respondents and the whole UKB cohort are compared. The major phenotypes are lifetime: depression (18%); panic disorder (4.0%); a specific eating disorder (2.8%); and bipolar affective disorder I (0.4%). All mental disorders are found less with older age and also seem to be related to selected social factors. In those participants who answered both MHQ1 (2016) and MHQ2 (2022), current mental health measure showed that fewer respondents have harmful alcohol use than in 2016 (relative risk 0.84), but current depression (RR 1.07) and anxiety (RR 0.98) have not fallen, as might have been expected given the relationship with age. We also compare lifetime concepts for test-retest reliability.

**Conclusions:** There are some drawbacks to UKB due to its lack of population representativeness, but where the research question does not depend on this, it offers exceptional resources that any researcher can apply to access. This paper has just scratched the surface of the results from MHQ2 and how this can be combined with other tranches of UKB data, but we predict it will enable many future discoveries about mental health and health in general.

## Background

Mental health disorders cause considerable suffering for a substantial portion of the population. There is still a lot about mental health and disorder that we do not understand, and there is an urgent need for research to improve our ability to promote good mental health or treat disordered mental health (1, 2). Early mortality of people with severe mental disorders from physical health conditions is likely part of a reciprocal relationship between poor physical and mental health, which reinforces the importance of considering mental health in wider medical research (3–5).

The UK Biobank (UKB) cohort study was established with the scale and depth to help researchers grapple with complex questions about conditions that are life-threatening or disabling in adults (6, 7). Such conditions seldom have a single cause; instead, their onsets and prognoses are associated with many risk factors, including genetic variants at multiple loci, exposures at any time from conception onwards, and interactions between them (8). Many conditions also tend to cluster, causing further problems in investigating specific causal mechanisms (9). These features of high complexity are true for most mental disorders (10–12). Traditional epidemiological designs of modest sample sizes and limited data capture are not well suited to these challenges (13). The very large sample size and detail of data in UKB offers great potential for investigating many facets of mental disorders in adults.

UKB recruited over half a million individuals aged 40-69 between 2006 and 2010. The baseline assessment was extensive, with physical measures, biological sample collection (blood and urine in all, saliva in 100,000), a touchscreen questionnaire and an interview that gathered information on early life and current behaviour, as detailed on their website (14). An impressive list of potential biomarkers has been established, and participants’ approximate addresses have been used to provide measures such as area deprivation, access to green spaces and air pollution (15). A subset of participants has been characterised even further through ‘enhancements’, including via multimodal imaging (including neuroimaging), activity/sleep monitoring, and cognitive testing. Further data, including information on health outcomes, have been obtained through consented linkage to routinely collected information, including hospital in-patient statistics, cancer registry, COVID-19 vaccination data, death certification, and data from the primary care electronic health records for a subset of the cohort (16).

To realise the potential of UKB for mental health research, and mental aspects of physical health, a mental health outcomes (MHO) consortium was created, which made recommendations for an online questionnaire about current and past mental disorders and associated features. In 2016-17 participants were invited via email and postal newsletter to complete this first web-based questionnaire (MHQ1), leading to responses from over 157,000 individuals by July 2017 (17). The core of the questionnaire was an assessment of lifetime depression and generalised anxiety status, which allowed genome-wide association studies (GWAS) for these disorders (18, 19), but also facilitated non-genetic studies, touching on many other aspects of mental health research, as detailed in a recent review of the use of UKB for mental health research (20).

Recognising the changeability of mental health and the limitations of a single questionnaire to capture complex diagnoses, the MHO consortium proposed a second mental health questionnaire (MHQ2). Alterations were made to reflect the changing priorities of research following the COVID-19 pandemic. Like its predecessor, MHQ2 was not a distinct entity, but a collection of ‘modules’ that contained measures on a particular diagnostic or exposure topic. When choosing measures, consideration was given to comparability and complementarity with other cohort studies in the UK (21) and elsewhere (22), and experiences of using MHQ1 data. Scales on current depression, anxiety and alcohol use were repeated from MHQ1 to allow examination of possible fluctuation of these symptoms. MHQ2 also aimed to:

- Enrich the lifetime depression phenotype including treatment response
- Investigate new disorders: panic disorder and eating disorders
- Update aspects of participants’ social circumstances.

## Methods

### Aim

This paper outlines the content of the MHQ2 questionnaire, describes the cohort, and gives an overview of participant responses. We compare responses to MHQ1 and MHQ2 to look for changes in the mental health of the cohort, as well as the consistency of measures that aim to report lifetime phenotypes.

### UK Biobank

UKB invited people aged 40-69 years living close to one of 22 assessment centres in England, Scotland, and Wales between 2006-2010 to take part, 503,309 (5.5%) agreed, and 91% remain in the cohort. Three main sources of data in UKB are: baseline assessment and samples; linkages such as hospital inpatient records; and ‘enhancements’ such as imaging and the MHQs. UKB data can be accessed by submitting a research protocol for approval. Over 4,000 research proposals had been approved by January 2024.

Linkage and enhancements have variable coverage and have been conducted at various timepoints, so caution is needed in the analysis and interpretation of such data, as shown in a recent timeline (20). Participants in UKB differ from the wider UK population by more likely being female, having higher educational attainment, and living in areas with less deprivation, though more urban than average (23). The ethnicity of participants is predominantly White British, with small numbers of participants from other ethnicities (24). The health and health behaviours of the UKB participants are better than average, with less smoking, lower incidence of cancer and lower mortality (23, 25).

### Questionnaire administration

UKB participants who had a valid email address on record were sent an invitation email in October to November 2022, which included a hyperlink to a personalised questionnaire. If there was no response, a reminder email was sent two weeks after the initial invitation. Respondents who started, but did not complete, the questionnaire were sent a reminder two weeks after they last accessed it. Finally, a second invitation was sent to non-responders two months after their previous one (around January 2023). All participants were also able to access the online questionnaire via the UKB participant website, as explained in annual postal newsletters (sent to people with no email address). Respondents were asked to enter their date of birth as an identity check, and the data for any respondents whose entered date of birth did not match the information previously provided are not released. We define someone as having completed the MHQ2 if they completed the mental health modules shown in Table 1 (there was also a COVID-19 module) and their MHQ2 data was available, which excludes any participants who failed the identity check or withdrew their data between answering the questionnaire and our data access. This means that ‘non-completed’ includes a few participants who have answered some or all of the modules.

**Table 1.**
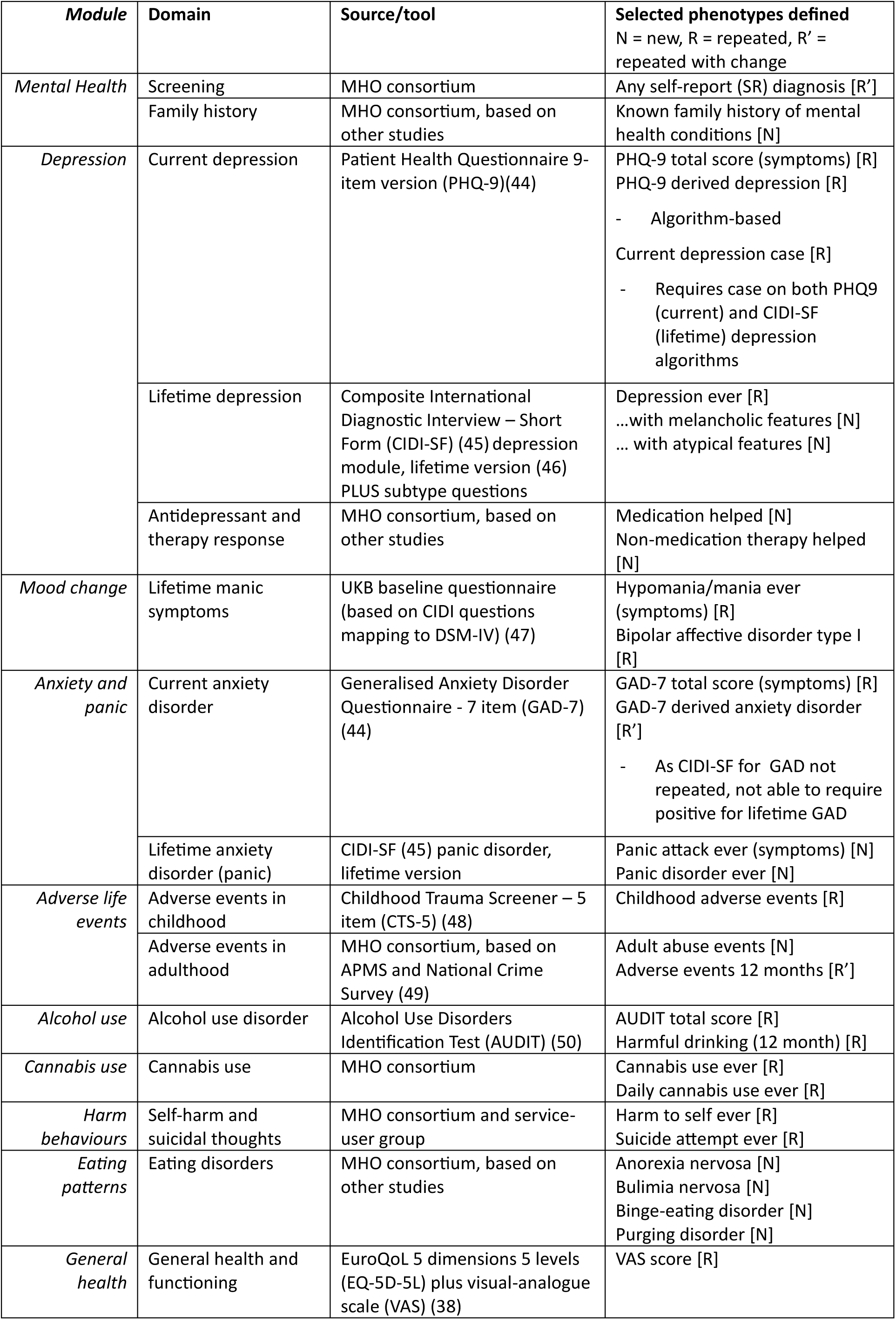
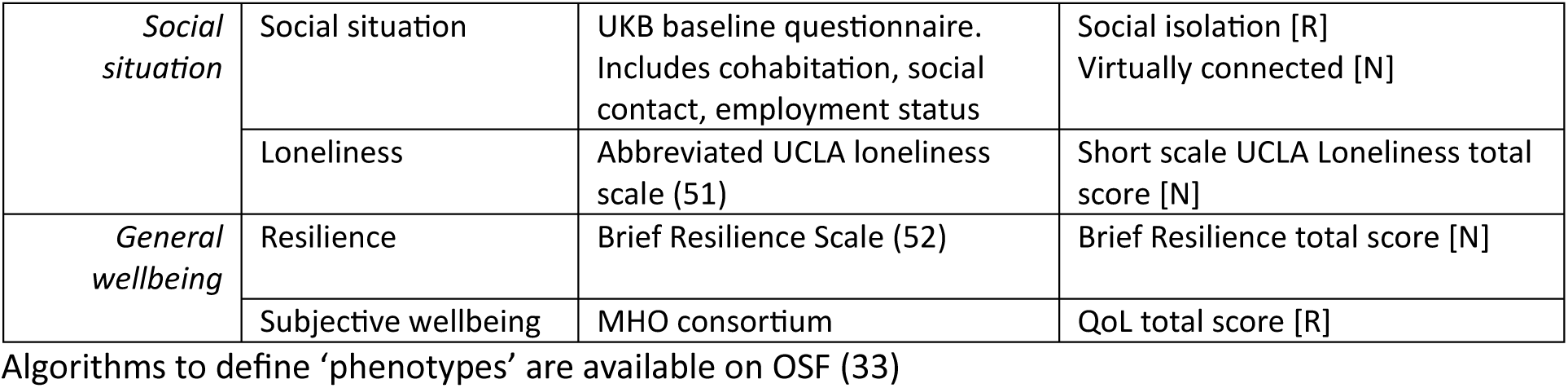
Modules of the MHQ2.

### Questionnaire development

The questionnaire domains were agreed upon by the consortium after a prioritisation survey and feedback from the Psychiatric Genomics Consortium (PGC) leads (26). Measures were discussed internally and with investigators who had experience with several other studies, including Genetic Links to Anxiety and Depression (GLAD) (27), The Scottish Health Research Register & Biobank (SHARE) (28), English Longitudinal Study of Ageing (ELSA) (29), the Eating Disorders Genetics Initiative (EDGI) (30), and the Adult Psychiatric Morbidity Survey (APMS, or Survey of Mental Health and Wellbeing) (31).

The measures chosen are provided in Figure 1, which shows how some modules were repeated exactly from MHQ1, and some were changed. For example, MHQ1 asks participants if they had been diagnosed with “panic attacks or panic disorder”, but in MHQ2, people are separately asked whether they have been diagnosed with “panic attacks” and then “panic disorder” to discriminate between them. Such changes mean some sections were not exactly equivalent, so were labelled ‘repeated with changes’ in Figure 1. To maximise the acceptability of the questionnaire length, some of the measures from MHQ1 were not repeated. These choices were made according to the relative support in the consortium for inclusion. The modules are also shown in Table 1, which shows in greater detail the measures used in each domain and the phenotypes that could be derived from these measures. A more detailed explanation is available on the UKB data showcase (32) or the OSF site for this project (33).

**Figure 1:**
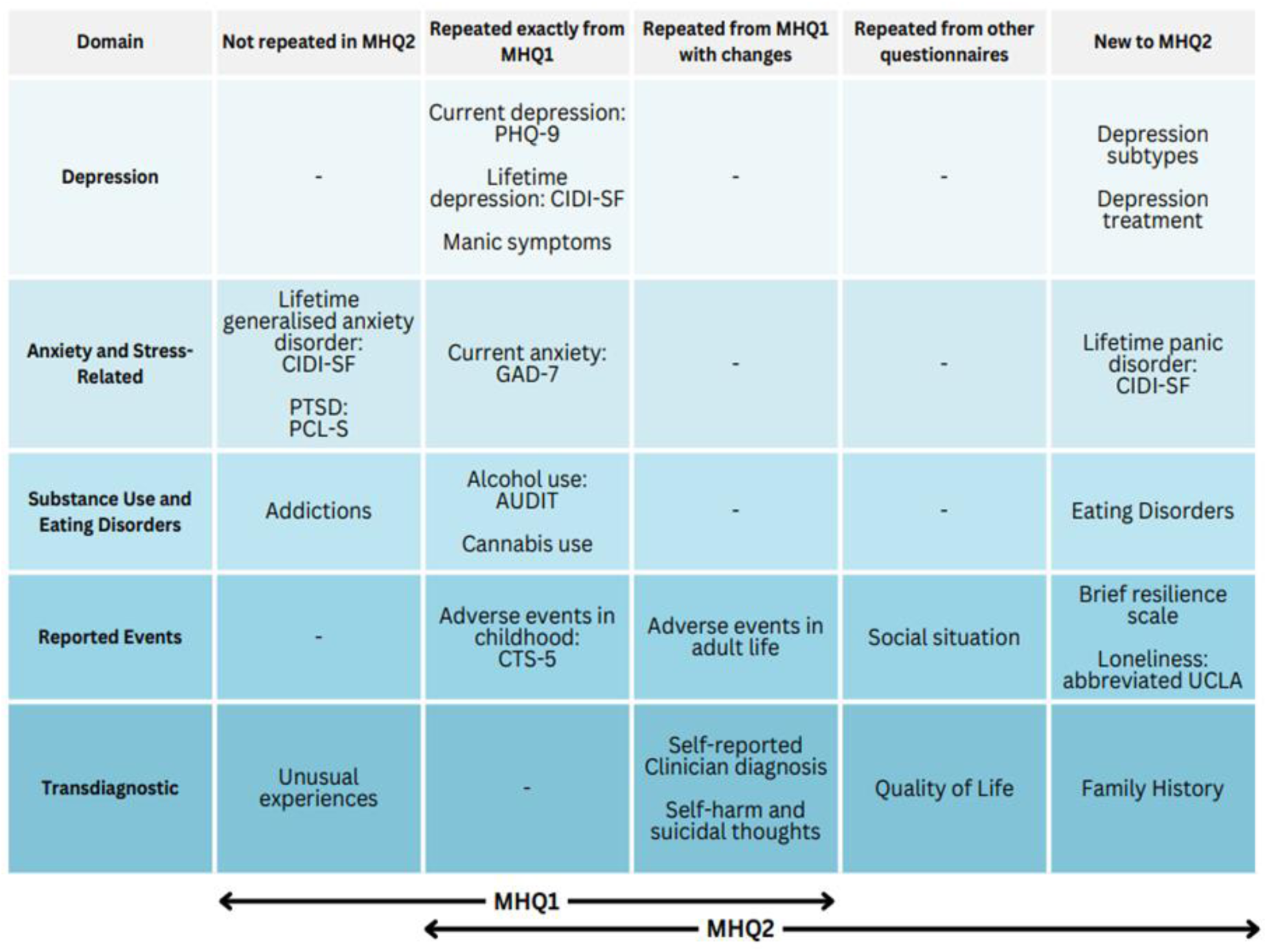
Summary of the topics included in MHQ1 and MHQ2, illustrating the overlap

### Algorithms and Code in R

Symptoms, questionnaire scores, experiences and (probable) diagnostic status that can be derived from the questionnaire are generically termed ‘phenotypes’ (with no assumption of genetic causation). For each module or group of modules, small groups of MHO members with expertise in those subjects produced algorithms to derive these phenotypes from questionnaire responses.

These were then checked and edited to create a consistent style and circulated to the whole consortium for comments.

A team of coders, including members of the MHO and junior colleagues, worked in small groups to render the algorithms for each module in R code, with a check by another coder outside of that module subgroup. Lastly, an over-arching script was created that brought the code from all the modules’ subgroups together, including a final few definitions that required responses from multiple modules. The algorithms and code for processing MHQ1 and MHQ2 are available on our OSF and GitHub pages (33, 34).

A clinical diagnosis is informed by diagnostic criteria, including the lack of alternative explanations and the presence of impact on the patient. However, when we use questionnaires, we are unable to take wider issues into account. The criteria we use for the “lifetime depression” phenotype, for example, are therefore referred to as quasi-diagnostic because they are informed by diagnostic criteria, but fail to fully examine a participant. When we refer to a group with “lifetime depression”, we mean only that they met the criteria for lifetime depression on our measure, not that they were fully assessed.

### Ethical permission

UKB operates inside the ethical framework of the UK Health Research Authority. Participants gave consent for their data to be used and can withdraw at any time, with an option to withdraw their existing data from future analysis. Research Ethics Committee opinion has been sought for UKB and each enhancement from the North West - Haydock Research Ethics Committee, including November 2022 (21/NW/0157, amendment 07).

The data in this paper were accessed through UKB-approved research number 82087, downloaded 23/10/2023, with withdrawals downloaded 11/06/2024 (except for the participation numbers, which were provided by UKB’s scientific team with a censoring date of 11/06/2024).

### Availability of data and materials

Data are available subject to UK Biobank procedures (14). Resources that the consortium has produced to help future researchers using MHQ2 data, including a document outlining recommended scoring and algorithms, and R code to facilitate implementation of those algorithms, can be found on the Open Science Framework (OSF) site for this project (33).

### Analysis

We planned a descriptive summary of the questionnaire respondents and responses, considering:

- Participants: Compare participants in the 2022 MHQ2 wave against the 2016 MHQ1 wave and the full original UKB cohort.
- Major phenotypes: Present numbers with lifetime mental health disorders in MHQ2 and compare personal characteristics between groups.
- Social factors: Present selected social data - social isolation, loneliness, resilience and health-related quality of life - stratified by lifetime mental disorder phenotypes.
- Current mental health: Describe and visualise current mental disorders for the 2022 MHQ2 wave against the 2016 MHQ1 wave.
- Test-retest reliability: Look at the consistency of lifetime disorder status between the 2022 wave and the 2016 wave.

For a comparison of ‘current’ mental health, we looked at the proportions of respondents meeting case criteria according to PHQ-9 (depression), GAD-7 (anxiety) and AUDIT (harmful alcohol use), restricting to those who answered in both waves. Age- and sex-specific proportions are given, with the age groups in seven-year blocks, which reflects the amount of time between the two waves. This was designed such that most participants moved one age block between MHQ1 and MHQ2, although this was not exact. Line graphs illustrate the distributions in the two waves. The relative risk of depression, anxiety and harmful alcohol use between 2016 and 2022 overall and in age and sex groups were calculated.

For a comparison of ‘lifetime’ status in depression ever (CIDI-SF lifetime), bipolar ever (adapted CIDI), self-harm ever (self-report), cannabis ever (self-report) and any clinician diagnosis (self-report), we report percentage agreement and Cohen’s kappa, with kappa being agreement corrected for agreement by chance (35).

Age at MHQ2 was defined for those who completed it as their age when completing the last module, similarly age at MHQ1 was age at completion. For those who did not complete the questionnaire (e.g., when using all UKB participants as a comparison group), we took the age they would have been at the median date of MHQ2 questionnaire completion. This includes the age someone would have been if they had not already died. We excluded any participants who had missing data in the variables used to define age (n=2). Except where stated, we include respondents of MHQ2 regardless of whether they participated in MHQ1.

We refer to the proportion of cases in the respondents. Due to the issues with representativeness, we do not attempt to make inferences about population prevalence, so hence do not include confidence intervals. Percentages are given in whole percentage points unless under 10% when one decimal place is used, as recommended for clarity (36). Where rare disorders and detailed breakdown of age or ethnicity intersected to make a cell size of less than 10 participants we have merged fields out of caution for confidentiality (37).

## Results

### Sample

Data provided by UKB’s scientific team showed that, after accounting for deaths and withdrawals, 457,653 participants were eligible to complete the MHQ2 (Figure 2). Of these, UKB was able to invite 329,902 (72%) participants by email, and 53% of those contacted answered at least one module of the questionnaire, alongside a small number (n=530) who accessed the questionnaire through their participant website. 175,266 participants completed at least one module with 169,252 completing all the mental health modules in the questionnaire (97% of those who started). The response dates cluster around early November 2022 (approx. 80%), when most emails went out, with a smaller peak in approximately January 2023 when reminder emails were sent. The data available to researchers may vary due to late completion, quarantine of questionnaires when the identity check was failed (approx. 0.3% of respondents) and participants withdrawing from UKB asking for no further analysis.

**Figure 2.**
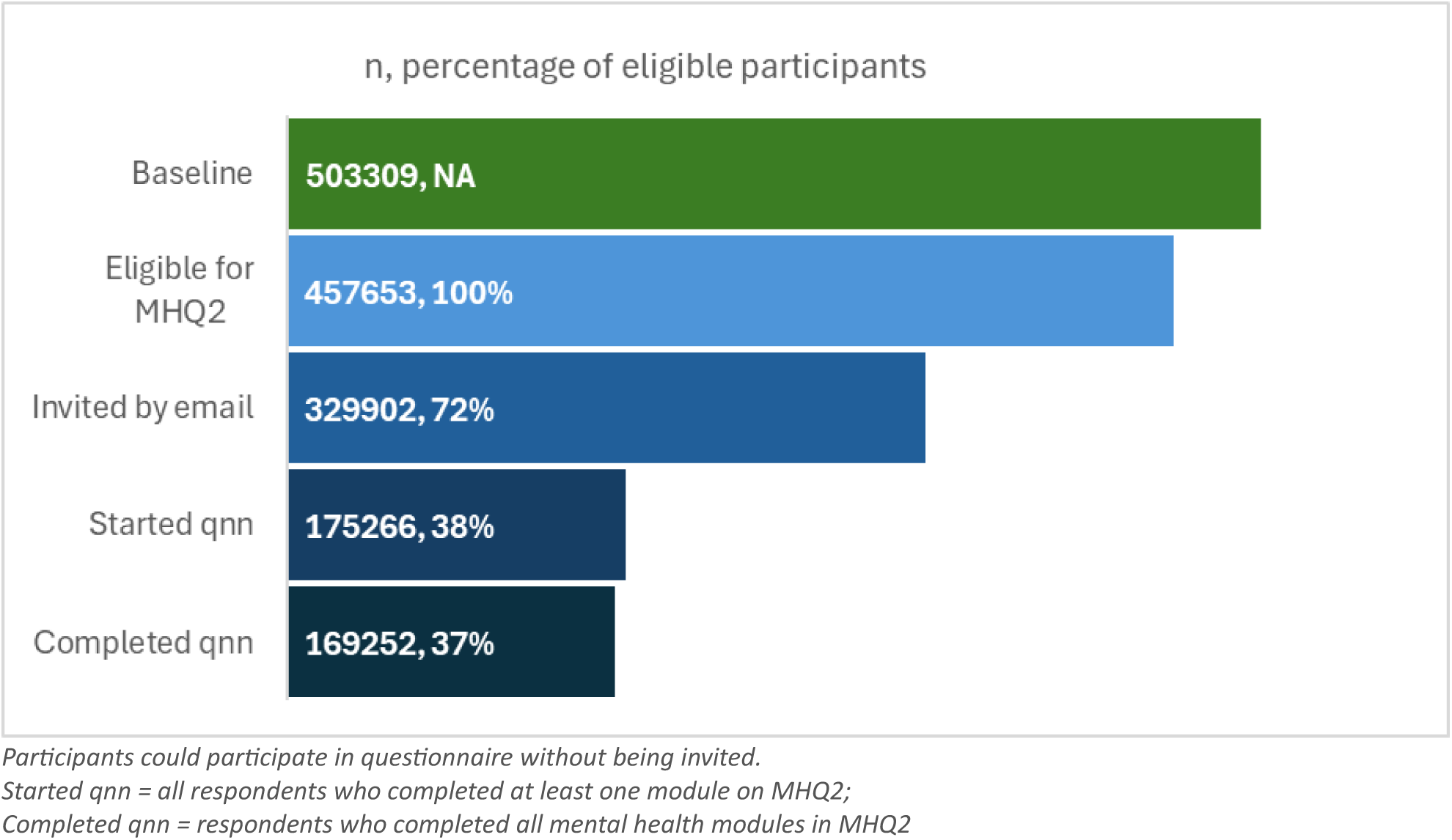
Number of UKB participants invited, starting and completing the MHQ2 questionnaire, as a proportion of those eligible for the questionnaire

Figure 3 shows that the 169,252 UKB participants who completed MHQ2 consisted of 111,272 who had also completed MHQ1 plus 57,980 who had not. This gives a sample of 111,272 who have longitudinal data (both waves), and a sample of 215,250 who have data for either MHQ.

**Figure 3.**
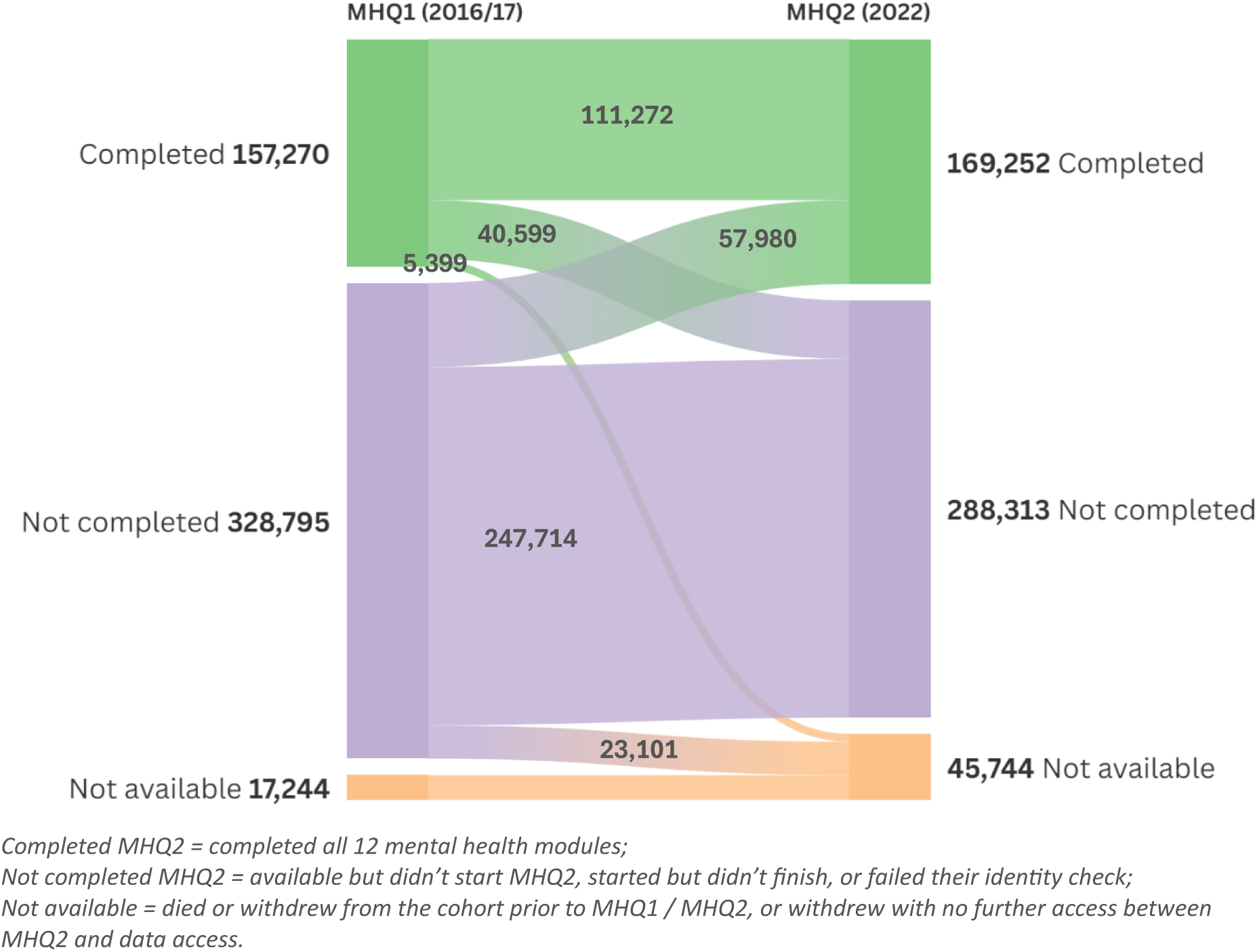
The flow of participants illustrating those who have data from completing the first and second mental health questionnaires, the cross-over and participants who have died or withdrawn.

### Sample Characteristics

Data available to researchers was of 169,253 respondents. The characteristics of the respondents in the MHQ2 wave are shown in Table 2. Compared to those for whom we have data from MHQ1, they are an average of five years older (after an interval between MHQ1 and 2 of approximately six years), and very similar in sex, ethnicity, and the distribution of selected social factors. The comparison of the MHQ2 wave with the UKB cohort showed that respondents are less likely to be deprived (4.6 percentage points less likely to live in a deprived area), report longstanding illness (5.6 percentage points less likely to have reported a longstanding illness) or be from a minority ethnic group (2.8 percentage points more likely to be White) than the cohort from which they are drawn. Table 2 shows the MHQ1/2 waves particularly under-represent people from Black and Asian ethnicities.

**Table 2.**
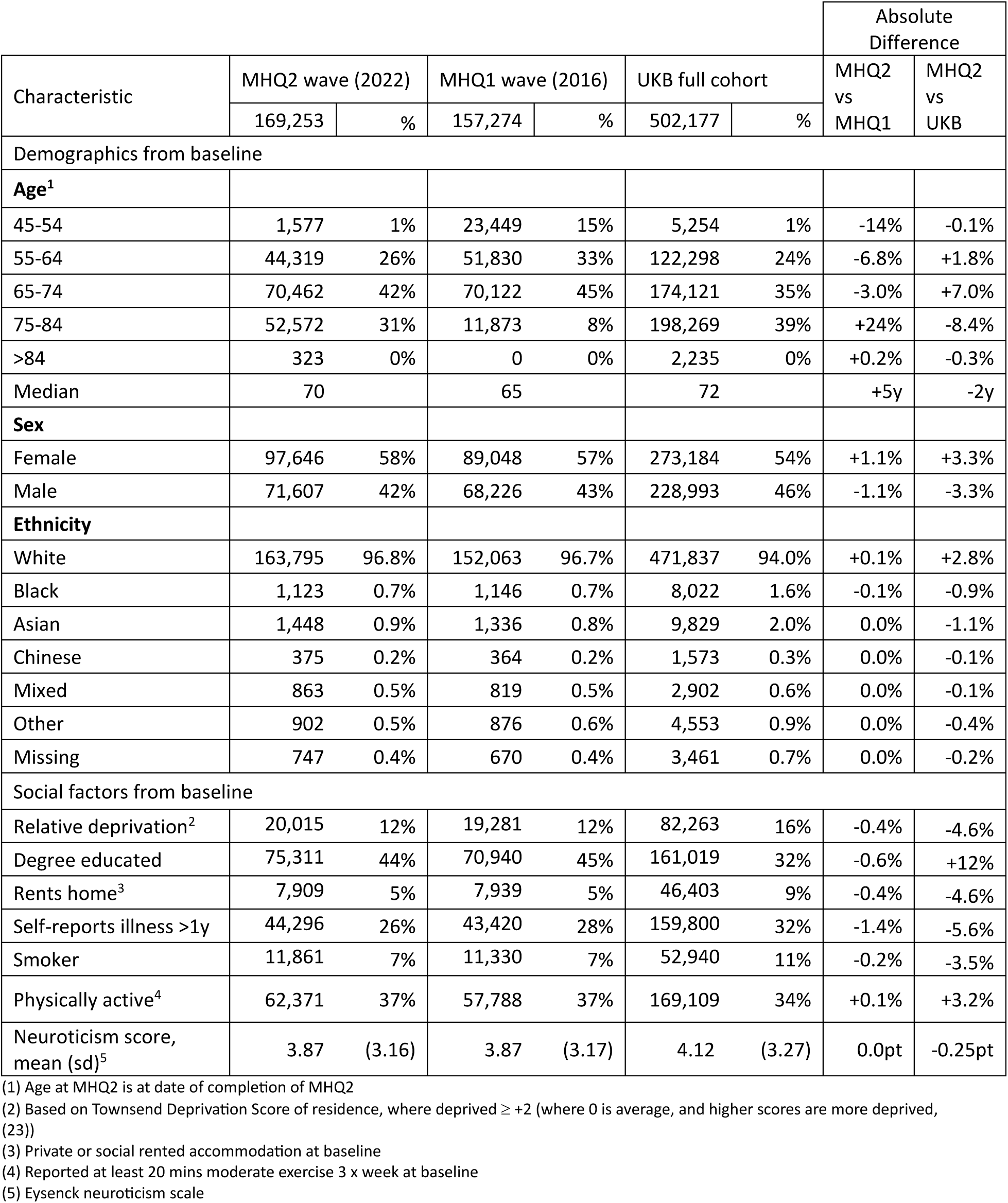
Characteristics of the respondents to each of the mental health questionnaires (available to analyse in 2024) with comparison to the UK Biobank cohort.

### Disorders

In Table 3 and the UPSET plot Supplementary Figure 1, participants are categorised by lifetime disorder status based on symptom-based definitions for depression, panic disorder, eating disorders and bipolar type I. Participants were most likely to be positive for lifetime depression (18%), followed by panic disorder (4.0%), any specific eating disorder (2.8%), and bipolar affective disorder type I (0.4%). Seventy-one percent of the MHQ2 respondents did not meet the criteria for any of the studied lifetime disorders.

**Table 3.**
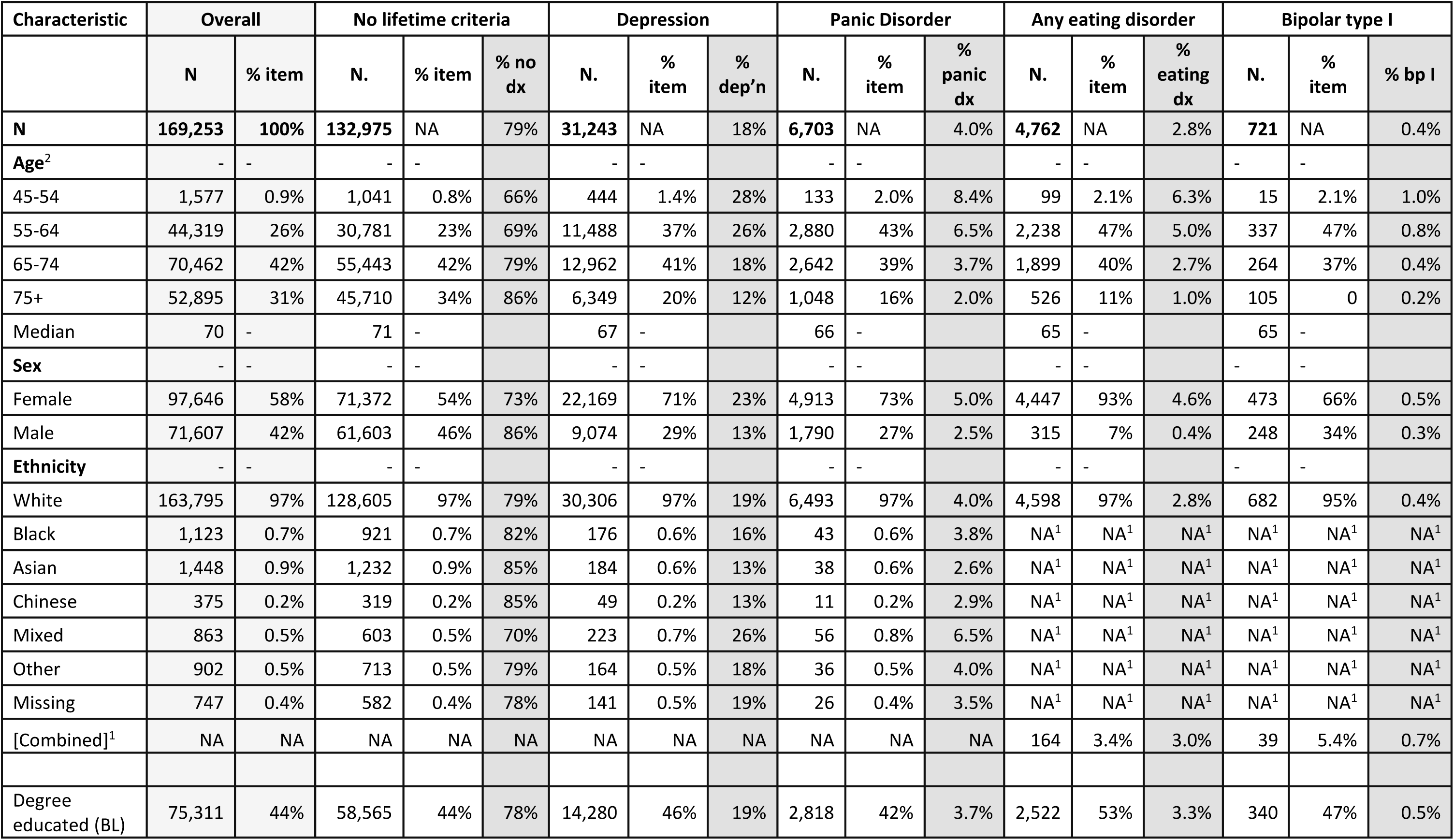
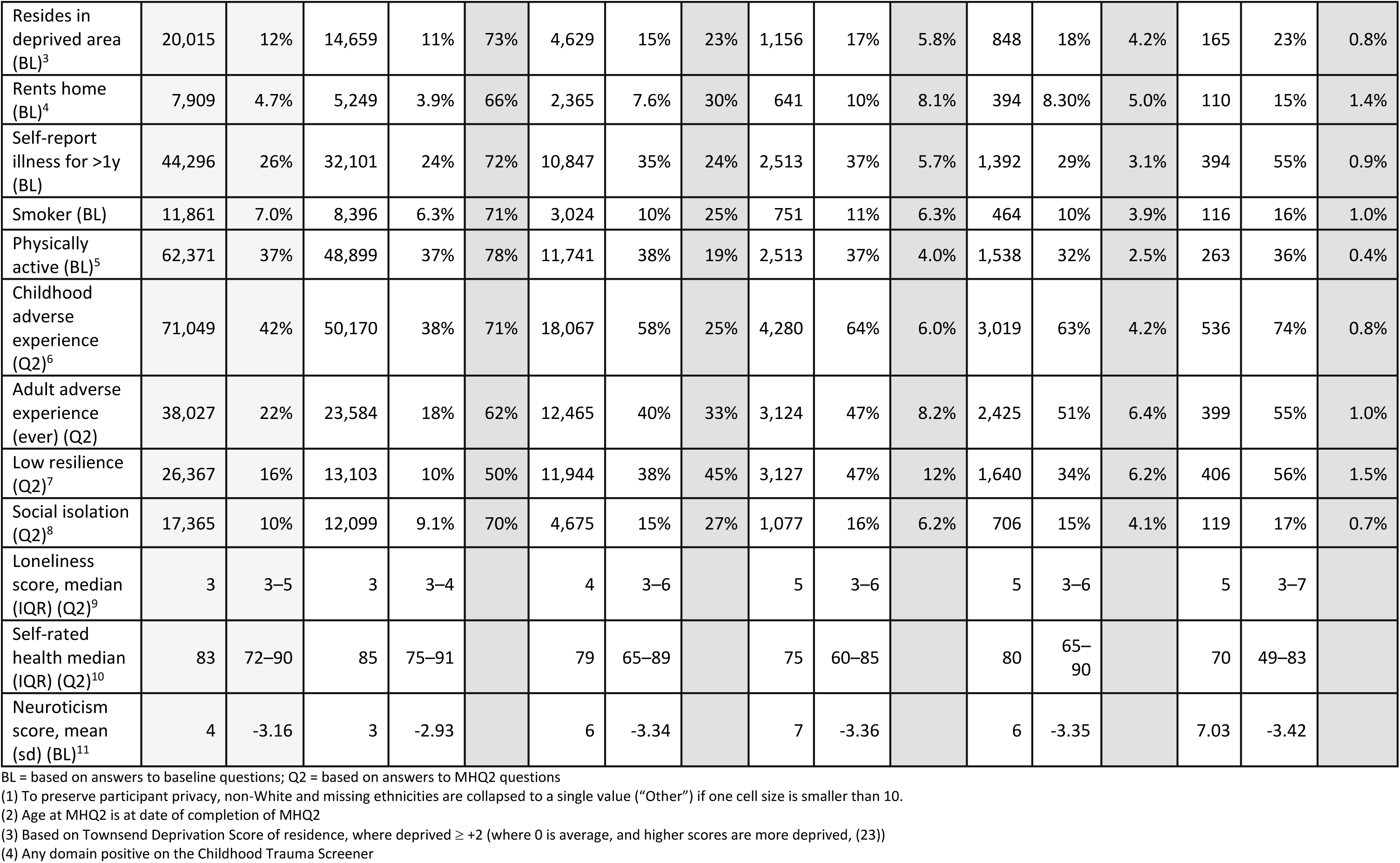
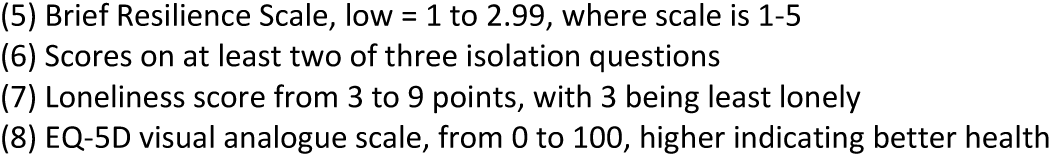
Characteristics of participants according to symptom-based criteria for lifetime disorders. Characteristics from baseline (BL) or MHQ2 (Q2).

The symptom-based definition of panic disorder was included for the first time in the MHQ2 wave. Self-reported clinician diagnosis of panic disorder was much lower (0.2%) than the symptom-based definition (4.0%), although 4.5% reported a clinician diagnosis of a panic *attack*. For context, in the MHQ1, the symptom-based definition of lifetime generalised anxiety disorder (GAD) was met by 7%.

Within the eating disorders category were four specific disorders, and respondents could fall into more than one in their lifetime. The most common eating disorder was anorexia nervosa (1.7% of respondents), followed by purging disorder (0.9%), bulimia nervosa (0.4%), and binge-eating disorder (0.2%). Self-report of an eating disorder diagnosis was somewhat rarer, for example, self-report of anorexia nervosa was 0.5% compared with 1.7% identified by the symptom-based definition.

The proportion with any lifetime disorder (symptom-based definition) is patterned by age, ethnicity and sex: higher in younger age (45-64: 34%) rather than old (75-84: 14%); higher in those of Mixed ethnicity at 30%, than White (21%) or Asian (15%); higher in female (27%) than male (14%). The increased proportion in female respondents was particularly marked in eating disorders (F: 4.6%, M: 0.4%).

Supplementary Figure 1 shows the degree of comorbidity between the phenotypes in Table 3 with the addition of self-harm ever. The lifetime self-harm phenotype includes 7765 (4.6%) participants, most (57%) also meeting the criteria for another phenotype, most commonly depression. Of those with lifetime depression, 29% also have one or more other phenotype, most commonly self-harm.

### Social Factors

The distributions of social factors are shown in Table 3, with the results in female participants in Supplementary Table 1 and male participants in Supplementary Table 2. The variables assessed at baseline were area-level deprivation, education (‘degree educated’), housing tenure (‘rents home’), longstanding illness, smoking, physical activity, and neuroticism – of which area-level deprivation, renting a home and smoking seemed to be particularly related to lifetime disorder. The group of respondents that met the criteria for eating disorders and panic disorder have similar characteristics to those respondents with depression and bipolar, although those with lifetime eating disorders are more likely to have a degree qualification (eating disorder 53%; depression 46%; no mental disorder 44%).

Adverse events in childhood were reported by 42% and adverse events in their adult life by 22%. In the group that met no symptom-based definition in the MHQ2, 38% reported a childhood event, increasing to 58% in those with lifetime depression, and rising further for rarer disorders – with the same pattern for events in adult life. The pattern of adverse childhood event report stratified by major phenotypes is the same in both men and women, Supplementary Tables 1 and 2.

Resilience was ‘low’ on the brief resilience scale in 10% of the group that met no symptom-based definition in the MHQ2 but 34% to 56% in those who met at least one symptom-based definition for a lifetime disorder. Those with at least one lifetime disorder had a slightly higher frequency of social isolation and loneliness than those who met no symptom-based definitions but were less marked than the pattern for low resilience.

Self-rated health was taken from the EQ-5D-5L visual analogue scale, where respondents rate their health ‘today’ using a slider from 0 to 100 (38). This rating was an average of 88 for people with no lifetime criteria, 80 for those with depression and eating disorder history, 75 for those with panic disorder and 70 for those with bipolar disorder. More than a quarter of those with lifetime bipolar disorder had marked their health as below 50 on the scale.

### Current Mental Health between 2016 (MHQ1) and 2022 (MHQ2)

Three measures of current mental health were included in both questionnaires: AUDIT for harmful alcohol use, PHQ-9 for depression, GAD-7 for generalised anxiety (AUDIT “in the last year”, PHQ-9 and GAD-7 “in the last two weeks”,). Restricting to people with data from both the MHQ1 and MHQ2 waves, Supplementary Tables 3 and Figures 4 - 6 show the comparison of proportions meeting the criteria, stratified by age and sex, using six-year age bands so that most respondents moved to the next age band between waves. Supplementary Table 4 shows the relative risk of meeting these criteria in 2022 versus 2016 in each age-sex group and overall. The figures show that, within each wave, the slope of proportion with depression, generalised anxiety and harmful alcohol use by age is downwards, indicating current mental disorder is less with increasing age. Between questionnaires, harmful alcohol use decreased along lines predicted for the older cohort (Figure 4), with a relative risk of harmful use for all respondents being 0.84 in 2022 compared to 2016. This does not occur for depression (Figure 5) or generalised anxiety (Figure 6), where the curve shifts, and the respondents overall have approximately the same risk of having current depression or anxiety in 2022 as they had in 2016 (relative risk depression = 1.07, anxiety = 0.98) despite the increasing age of respondents.

**Figure 4.**
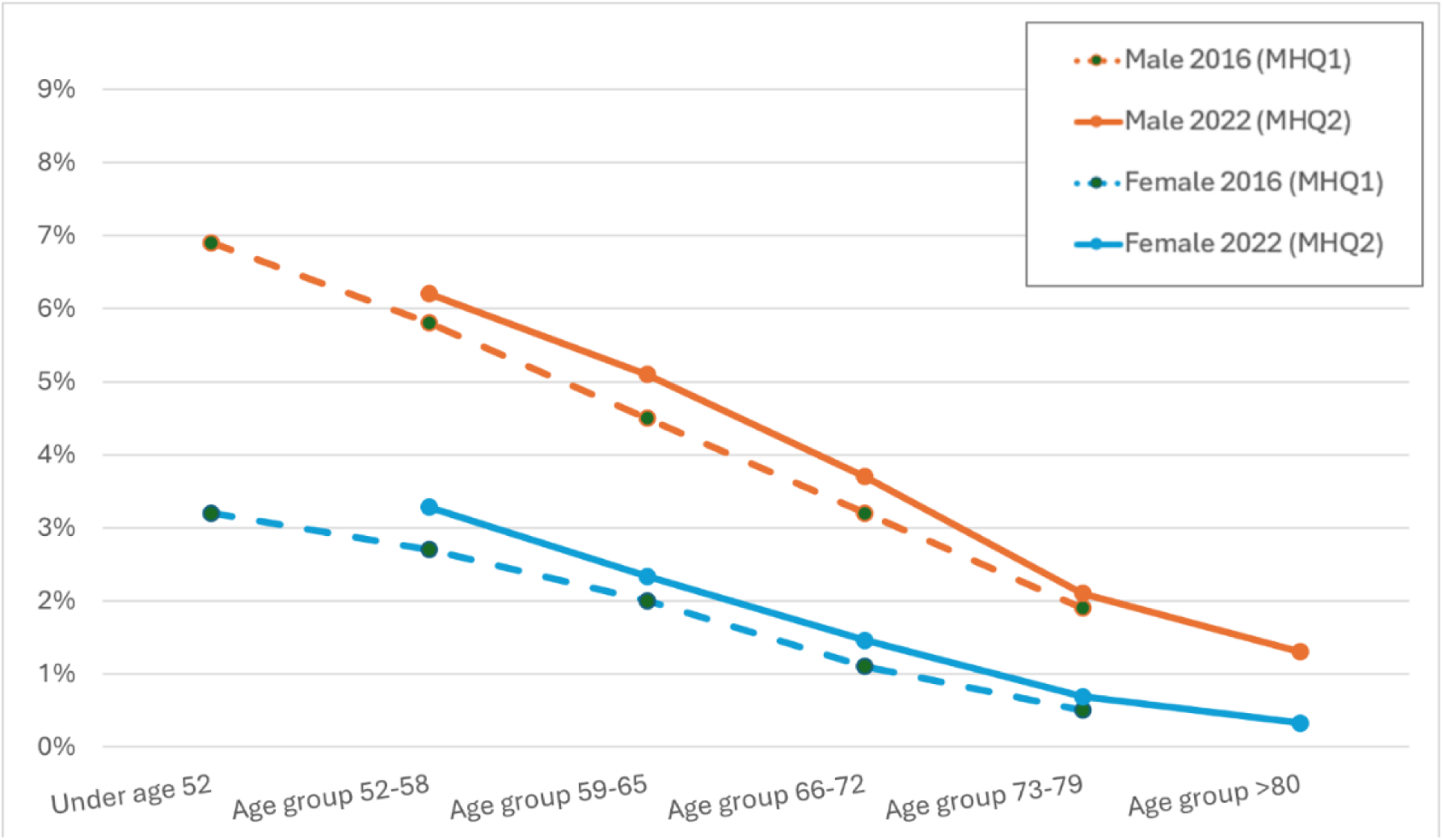
Percentage in each age group depressed according to ‘PHQ-9-derived current depression’ outcome by sex and age at completing questionnaires in 2016 and 2022

**Figure 5.**
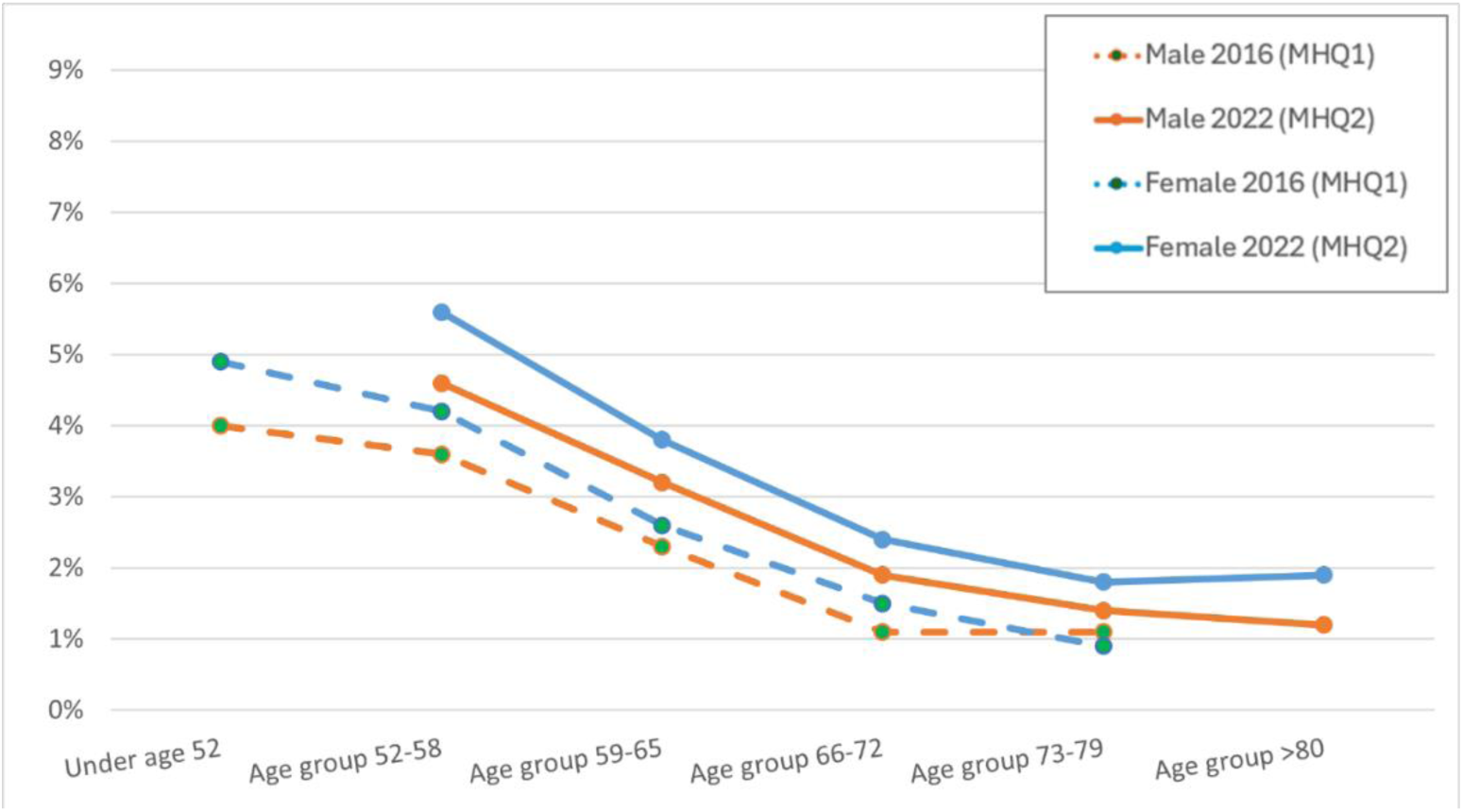
Percentage in each age group having generalised anxiety disorder according to ‘GAD-7- derived anxiety’ by sex and age at completing questionnaires in 2016 and 2022

**Figure 6.**
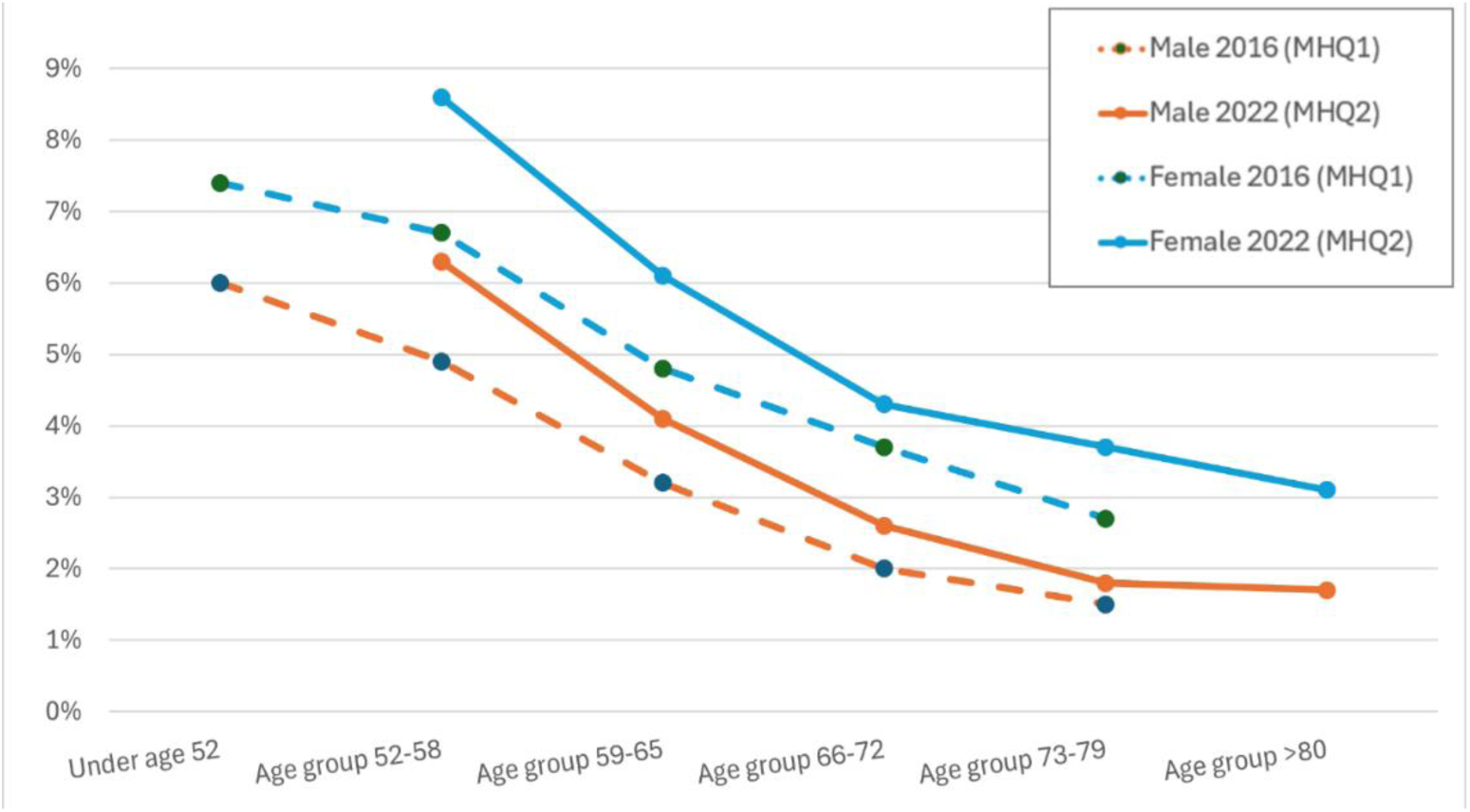
Percentage in each age group with harmful alcohol use according to AUDIT outcome by sex and age at completing questionnaires in 2016 and 2022

The largest relative risk increase in age and sex-stratified groups was in the women aged 73-79 category, with double the risk of depression, a 40% higher risk of generalised anxiety and harmful alcohol use, although absolute risk remained fairly low (depression = 1.8%, anxiety = 3.7%, alcohol = 0.7%).

Table 4 shows the difference in results from measures that report lifetime phenotypes: two symptom-based definitions of lifetime disorder (depression and bipolar), two lifetime behaviours (self-harm and cannabis use), and self-report of any clinician diagnosis, in those who completed both MHQ1 and MHQ2. Across these phenotypes, people were more likely to meet the criteria in MHQ1 than in MHQ2, except for self-harm. Kappa values show poorer agreement for the two symptom-based definitions (depression 0.53, bipolar 0.30) compared with the others (any clinician diagnosis 0.66, self-harm 0.67, cannabis 0.81). The lack of agreement between the questionnaire waves may in part be because of new-onset cases that started after completing MHQ1. However, after taking into account onsets after MHQ1 (including unknown onsets), between 75% to 93% of the test-retest variation in direction ‘Met 2 not 1’ and all in direction ‘Met 1 not 2’ remains unexplained.

**Table 4.**
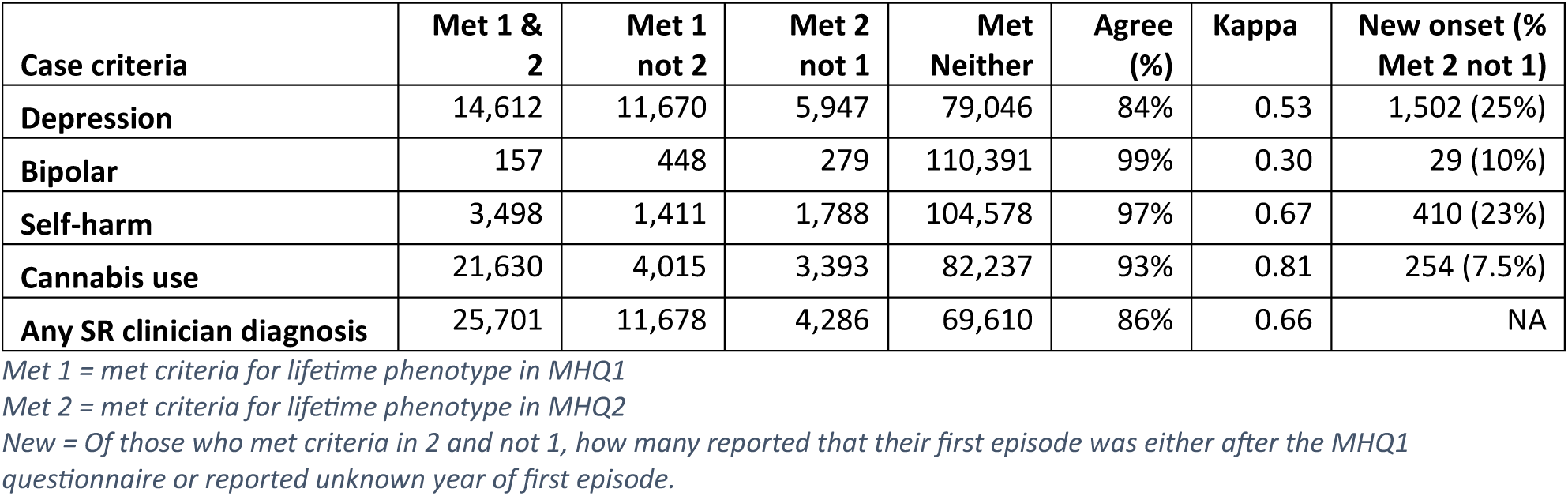
Comparison of lifetime phenotype status in MHQ1 and MHQ2 in those participants who completed both questionnaires (n=111,275)

| **Case criteria** | **Met 1 & 2** | **Met 1 not 2** | **Met 2 not 1** | **Met Neither** | **Agree (%)** | **Kappa** | **New onset (% Met 2 not 1)** |
| --- | --- | --- | --- | --- | --- | --- | --- |
| **Depression** | 14,612 | 11,670 | 5,947 | 79,046 | 84% | 0.53 | 1,502 (25%) |
| **Bipolar** | 157 | 448 | 279 | 110,391 | 99% | 0.30 | 29 (10%) |
| **Self-harm** | 3,498 | 1,411 | 1,788 | 104,578 | 97% | 0.67 | 410 (23%) |
| **Cannabis use** | 21,630 | 4,015 | 3,393 | 82,237 | 93% | 0.81 | 254 (7.5%) |
| **Any SR clinician diagnosis** | 25,701 | 11,678 | 4,286 | 69,610 | 86% | 0.66 | NA |

*Met 1 = met criteria for lifetime phenotype in MHQ1*
*Met 2 = met criteria for lifetime phenotype in MHQ2*
*New = Of those who met criteria in 2 and not 1, how many reported that their first episode was either after the MHQ1 questionnaire or reported unknown year of first episode.*

Supplementary Table 5 and 6 show the sex-stratified results, and they are similar; for example, self-harm reporting in women had a kappa of 68%, and in men 65%.

## Discussion

This paper introduced UK Biobank’s second mental health questionnaire. The original MHQ was one of the largest mental health surveys ever reported. This second wave gives an even larger sample size, more detail, and coverage of broader aspects of mental health. 169,253 UK Biobank participants completed the MHQ2, which brings to 215,252 the number who have at least one wave of the mental health questionnaire that can be analysed. We have selected a few notable features to describe, which we hope will stimulate interest in this resource. These were: the characteristics of MHQ2 wave participants; lifetime disorders categories; and selected social factors. We also explored changes in current disorder status and agreement of lifetime phenotypes between wave MHQ1 and MHQ2.

There was a substantial overlap of participants who completed the previous wave, and participant characteristics of the two waves were similar. As shown in the previous paper (17) the biases in completion of the mental health questionnaires resembled and amplified the biases in recruitment to UKB. The respondents to MHQ2 were less likely to be deprived or have a chronic illness than the overall UKB cohort, and 97% were from a White ethnic group.

Depression was the most common lifetime disorder at 18% in the MHQ2 wave, although this is less than the 24% reported in the MHQ1 wave. Next in this questionnaire was panic disorder (4.0%) and eating disorder (2.8%) with bipolar type I (0.4%) the least common. All four disorders have similar findings in social and health characteristics: increased adverse child events; more difficult and deprived personal circumstances; and poorer perceived health. This pattern is more pronounced with rarer disorders, such that deprivation and self-rated health are least adverse in the group with depression, worst in the group with bipolar, and the groups with panic disorder or eating disorders are in-between.

The finding that those with lifetime disorders score worse on self-rated health is unsurprising, as health may be poorer both because of the mental disorders themselves and the greater risk of developing physical illness. Conversely, people with physical disorders are at higher risk of developing a mental disorder (9). UK Biobank is a valuable data source for studying these issues, and repeated measures to give longitudinal data will help. For instance, four questions from the PHQ-9 were asked at baseline and have been repeated in several enhancements (sleep, cognitive testing, imaging visits, MHQ1 and MHQ2) and the EQ-5D-5L (health-related quality of life) reported here from MHQ2 was also asked in the UKB pain questionnaire in 2019, and is due to be repeated.

### Repeated measures

In the MHQ1 wave, both lifetime and current mental health disorders appeared to be rarer in old age. This could be an age effect (mental health gets better as people age) or a cohort effect (people born earlier have better mental health). Using repeated current mental disorder scales, we found that the frequency of alcohol use disorder had decreased over six years, suggesting an age effect, but depression and anxiety levels had stayed fairly unchanged as the cohort aged, suggesting a cohort effect. Another possibility is that 2022 was an era of higher anxiety and depression due to the COVID pandemic and knock-on effects, for example, prolonged health service disruption, which may make it difficult for any age-related improvement in mental health to be demonstrated at this time. The short-term deviation of mental distress from the COVID pandemic has been well-documented (for example,(39)), but the long-term mental health outcomes of the period are still unknown (40), and may merit monitoring.

We investigated the test-retest consistency for phenotypes that we would expect to be similar between waves: lifetime depression, bipolar affective disorder type I, self-harm, cannabis use and self-report of any clinician diagnosis. In all cases, there was high agreement on phenotype status (from 84% for depression to 99% for bipolar), although kappa (which corrects for agreement by chance) revealed that lifetime depression and lifetime bipolar statuses were somewhat inconsistent (0.53 for depression, 0.30 for bipolar), and this was not accounted for by new-onset. To meet criteria for depression and bipolar, several criteria needed to be met, frequently hinging on a dichotomous scoring of a Likert-like scale. For instance: “How much did these problems interfere with your life or activities? – A lot / Somewhat / A little / Not at all”. To meet the criteria for lifetime depression participants needed to endorse “A lot” or “Somewhat”. We can speculate that a person’s appraisal could change from “Somewhat” to “A little” over time, perhaps to a more positive interpretation as they age. Therefore, a change from meeting criteria may occur with only modest changes in symptom reporting. To meet the criteria for bipolar, participants had to meet criteria for both depression and mania. The definition for ever self-harm and cannabis consisted of a few yes/no questions. For yes/no questions, and when talking about behaviours rather than feelings, there would be less reappraisal. It is not clear why the recall of self-report clinician diagnosis might be inconsistent, perhaps it hinges on recall of whether a disorder was diagnosed, rather than merely discussed or mentioned. These inconsistencies may complicate the interpretation of some of these outcomes, but it is possible they reflect the underlying uncertain boundaries between health and disorder (41, 42).

In this investigation, we also noted those who told us their disorder did not start before MHQ1 (i.e. new or unknown onset), which included 1,502 people with depression (4.7% of those with depression in MHQ1 or 2, or 1.6% of respondents) and 410 people who have self-harmed (6.1% of those with self-harm in MHQ1 or 2, or 0.4% of respondents). These are a small group, but those with late-onset mental disorders are a group that is potentially important to consider in this cohort of ageing.

### Strengths and limitations

UK Biobank is a valuable resource for investigating mental health, but it also presents challenges. The cohort consists of a convenience sample, and the non-representativeness observed at baseline assessments is exacerbated in the voluntary enhancements (such as the MHQs). The multiple enhancements with different samples and timings can complicate planning and interpreting analyses. Because of non-representativeness, researchers should not make population inferences based solely on UK Biobank data. We have not tested what effect the non-representativeness of the sample has had on our ascertainment of mental disorders and the pattern with social factors.

We have designed the UKB MHQ1 and MHQ2 with researchers familiar with large cohorts and clinical academics – leading to the use of widely accepted measures and clinically relevant outcomes. The completion of the questionnaire was also high once started, assisted by reminder emails. However, the MHQs are reliant on self-report, frequently of an episode of psychopathology that may have been many years ago. This may result in some inconsistencies, such as seen in the test-retest statistics – which we suggest is due to inconsistent recall and indistinct boundaries in mental disorders, which have no diagnostic tests.

Our approach to developing this paper and facilitating further research using MHQ2 has incorporated quality assurance processes when developing algorithms and code, which we are sharing with the wider research community. We enhance transparency by sharing resources, however, errors are still inevitable in such projects (43) so we encourage caution when using these resources, and feedback from others if they suspect any problems. This paper is just the beginning, which we hope will encourage other researchers to investigate more deeply. Further advice for people considering using UKB for mental health research is available in a recent review paper (20).

## Conclusion

The mental health questionnaires enhance UKB with large amounts of data on common mental disorders (particularly depression), and also on disorders that have not received much research in an older population, such as panic disorder and eating disorders, and update social factors a decade after baseline. Importantly, the UKB now has included transdiagnostic symptoms (self-harm, body weight/shape preoccupation, psychotic experiences) and psychological constructs (resilience, loneliness and neuroticism) that are infrequently or never available from clinical data. Repeated measures for items such as depressive symptoms and health-related quality of life, along with linkage to routine health data for follow-up, allow longitudinal and causal analyses. UKB offers a large sample size, extensive genetic information, biomarkers and imaging data, alongside detailed mental health data, which we hope will be used to help us understand more about mental health, and how we might prevent and treat poor mental health in the future.

## Supporting information

Supplementary Materials (separate file)

## Data Availability

Data are available subject to UK Biobank procedures https://www.ukbiobank.ac.uk/enable-your-research. Resources developed alongside this paper are available on the Open Science Framework (OSF) site for this project https://osf.io/c65t7/

## Abbreviations

AUDIT: Alcohol Use Disorder Identification Tool
CIDI-SF: Composite International Diagnostic Interview Short Form
DSM-IV: Diagnostic and Statistical Manual of Mental Disorders 4^th^ ed.
EQ-5D-5L: EuroQol questionnaire of 5 dimensions and 5 levels, plus VAS
GAD-7: Generalised Anxiety Disorder scale (for current GAD)
MHO: Mental Health Outcomes [consortium]
MHQ: Mental health questionnaire
OSF: Open Science Framework
PHQ-9: Patient Health Questionnaire (for current depression)
R: Statistical software
UKB: UK Biobank
VAS: Visual Analogue Scale (used in EQ-5D-5L)

## Declarations

### Ethics approval and consent to participants

UK Biobank has received favourable ethical opinion from North West - Haydock Research Ethics Committee 21/NW/0157, and this project has received approval from the UK Biobank access committee. All participants consented to participate.

### Consent for publication

Not applicable

### Availability of data and materials

Data are available subject to UK Biobank procedures https://www.ukbiobank.ac.uk/enable-your-research (14). Resources that the consortium has produced to help future researchers using MHQ2 data, including a document outlining recommended scoring and algorithms, and R code to facilitate implementation of those algorithms, can be found on the Open Science Framework (OSF) site for this project https://osf.io/c65t7 (33).

### Authors’ contributions

Conceptualisation: KASD, AD, AM, AJ, MR, EF, FS, GB, JH, RM, SO, TE, WL and MH.

Data curation: JC, MA, CH, HD, AK, DL, JM, RW, ZY and JZ.

Formal analysis: KASD, JC and KD.

Writing – original draft preparation: KD, JC and MH.

Writing – review and editing: All authors.

## Acknowledgements

This paper represents independent research funded by the NIHR Maudsley Biomedical Research Centre at South London and Maudsley NHS Foundation Trust and King’s College London. The views expressed are those of the author(s) and not necessarily those of the NIHR or the Department of Health and Social Care

## Supplementary Materials (separate file)

ST1: Characteristics of people meeting criteria for mental disorder **Female only**

ST2: Characteristics of people meeting criteria for mental disorder **Male only**

ST3: Age- and sex-stratified results for cases of depression, anxiety and harmful alcohol use in those who completed both questionnaires according to measures of current mental health

ST4: Relative risks of being a case on current mental health outcomes in 2022 compared to 2016 in age* and sex specific groups in 2022 in people who completed both MHQs. See ST3 for sample size by age and sex.

ST5 Comparison of lifetime conditions by participants, based on meeting criteria in MHQ1 and MHQ2, **Female only**

ST6 Comparison of lifetime conditions by participants, based on meeting criteria in MHQ1 and MHQ2, **Male only**

SFigure 1 Occurrence and co-occurrence of the studied mental disorder categories with the ten most common intersections shown

## References

1. GBD 2019 Mental Disorders Collaborators. Global, regional, and national burden of 12 mental disorders in 204 countries and territories, 1990–2019: a systematic analysis for the Global Burden of Disease Study 2019. The Lancet Psychiatry. 2022;9(2):137-50.

2. Arango C, Díaz-Caneja CM, McGorry PD, Rapoport J, Sommer IE, Vorstman JA, et al. Preventive strategies for mental health. The Lancet Psychiatry. 2018;5(7):591–604.

3. Prince M, Patel V, Saxena S, Maj M, Maselko J, Phillips MR, et al. No health without mental health. The lancet. 2007;370(9590):859-77.

4. Firth J, Siddiqi N, Koyanagi A, Siskind D, Rosenbaum S, Galletly C, et al. The Lancet Psychiatry Commission: a blueprint for protecting physical health in people with mental illness. The Lancet Psychiatry. 2019;6(8):675–712.

5. Ohrnberger J, Fichera E, Sutton M. The dynamics of physical and mental health in the older population. The Journal of the Economics of Ageing. 2017;9:52–62.

6. Sudlow C, Gallacher J, Allen N, Beral V, Burton P, Danesh J, et al. UK Biobank: An Open Access Resource for Identifying the Causes of a Wide Range of Complex Diseases of Middle and Old Age. PLOS Medicine. 2015;12(3):e1001779.

7. Allen NE, Lacey B, Lawlor DA, Pell JP, Gallacher J, Liam S, et al. Prospective study design and data analysis in UK Biobank. Science Translational Medicine. 2024;16(729).

8. Wu H, Eckhardt CM, Baccarelli AA. Molecular mechanisms of environmental exposures and human disease. Nature Reviews Genetics. 2023;24(5):332–44.

9. Ronaldson A, Arias de la Torre J, Prina M, Armstrong D, Das-Munshi J, Hatch S, et al. Associations between physical multimorbidity patterns and common mental health disorders in middle-aged adults: A prospective analysis using data from the UK Biobank. Lancet Reg Health Eur. 2021;8:100149.

10. Martins J, Yusupov N, Binder EB, Brückl TM, Czamara D. Early adversity as the prototype gene × environment interaction in mental disorders? Pharmacology Biochemistry and Behavior. 2022;215:173371.

11. Consortium TB, Anttila V, Bulik-Sullivan B, Finucane HK, Walters RK, Bras J, et al. Analysis of shared heritability in common disorders of the brain. Science. 2018;360(6395):eaap8757.

12. Uher R, Zwicker A. Etiology in psychiatry: embracing the reality of poly-gene-environmental causation of mental illness. World Psychiatry. 2017;16(2):121–9.

13. Burton PR, Hansell AL, Fortier I, Manolio TA, Khoury MJ, Little J, et al. Size matters: just how big is BIG?: Quantifying realistic sample size requirements for human genome epidemiology. International Journal of Epidemiology. 2008;38(1):263–73.

14. UK Biobank Website [Available from: www.ukbiobank.ac.uk.

15. Mamouei M, Zhu Y, Nazarzadeh M, Hassaine A, Salimi-Khorshidi G, Cai Y, et al. Investigating the association of environmental exposures and all-cause mortality in the UK Biobank using sparse principal component analysis. Scientific Reports. 2022;12(1):9239.

16. UK Biobank. Health Outcomes Overview v.2. https://biobank.ndph.ox.ac.uk/showcase/refer.cgi?id=596; 2024. Contract No.: Resource 596.

17. Davis KAS, Coleman JRI, Adams M, Allen N, Breen G, Cullen B, et al. Mental health in UK Biobank – development, implementation and results from an online questionnaire completed by 157 366 participants: a reanalysis. BJPsych Open. 2020;6(2):e18.

18. Coleman JRI, Peyrot WJ, Purves KL, Davis KAS, Rayner C, Choi SW, et al. Genome-wide gene-environment analyses of major depressive disorder and reported lifetime traumatic experiences in UK Biobank. Molecular Psychiatry. 2020;25(7):1430–46.

19. Purves KL, Coleman JR, Meier SM, Rayner C, Davis KA, Cheesman R, et al. A major role for common genetic variation in anxiety disorders. Molecular psychiatry. 2020;25(12):3292–303.

20. Davis KAS, Mirza L, Clark SR, Coleman JRI, Kassam AS, Mills NT, et al. The role of UK Biobank in mental health research: a review and commentary. OSF Preprints https://doiorg/1031219/osfio/bsywk. 2024.

21. Catalogue of Mental Health Measures. www.cataloguementalhealth.ac.uk2024 [Available from: www.cataloguementalhealth.ac.uk.

22. Psychiatric Genomics Consortium. About Us https://pgc.unc.edu/about-us/2024 [Available from: https://pgc.unc.edu/about-us/.

23. Fry A, Littlejohns TJ, Sudlow C, Doherty N, Adamska L, Sprosen T, et al. Comparison of sociodemographic and health-related characteristics of UK Biobank participants with those of the general population. American journal of epidemiology. 2017;186(9):1026–34.

24. Carress H, Lawson DJ, Elhaik E. Population genetic considerations for using biobanks as international resources in the pandemic era and beyond. BMC Genomics. 2021;22(1):351.

25. Schoeler T, Speed D, Porcu E, Pirastu N, Pingault J-B, Kutalik Z. Participation bias in the UK Biobank distorts genetic associations and downstream analyses. Nature Human Behaviour. 2023;7(7):1216–27.

26. Sullivan PF, Agrawal A, Bulik CM, Andreassen OA, Børglum AD, Breen G, et al. Psychiatric Genomics: An Update and an Agenda. American Journal of Psychiatry. 2018;175(1):15–27.

27. Davies MR, Kalsi G, Armour C, Jones IR, McIntosh AM, Smith DJ, et al. The Genetic Links to Anxiety and Depression (GLAD) Study: Online recruitment into the largest recontactable study of depression and anxiety. Behaviour Research and Therapy. 2019;123:103503.

28. Iveson MH, Ball EL, Doherty J, Pugh C, Vashishta S, Palmer CNA, et al. Cohort profile: The Scottish SHARE Mental Health (SHARE-MH) cohort – linkable survey, genetic and routinely collected data for mental health research. BMJ Open. 2024;14(1):e078246.

29. closer. English Longitudinal Study of Ageing 2024 [Available from: https://closer.ac.uk/study/english-longitudinal-study-of-ageing/.

30. Bulik CM, Thornton LM, Parker R, Kennedy H, Baker JH, MacDermod C, et al. The eating disorders genetics initiative (EDGI): study protocol. BMC psychiatry. 2021;21:1–9.

31. Catalogue of Mental Health Measures. Adult Psychiatric Morbidity Survey (APMS) 2019 [Available from: https://www.cataloguementalhealth.ac.uk/?content=study&studyid=APMS.

32. UK Biobank. Mental well-being web questionnaire 2023 [Available from: https://biobank.ndph.ox.ac.uk/showcase/refer.cgi?id=2800.

33. Davis KAS, Coleman JRI, UKB Mental Health Outcomes Consortium. UKB 2nd Mental Health Questionnaire Open Science Framework Home 2024 [Available from: https://osf.io/c65t7/.

34. Coleman J. R code for UKB MHQ2 algorithms github2024 [Available from: https://github.com/ColemanResearchGroup/MHQ2.

35. Byrt T, Bishop J, Carlin JB. Bias, prevalence and kappa. Journal of Clinical Epidemiology. 1993;46(5):423–9.

36. Cole TJ. Too many digits: the presentation of numerical data. Arch Dis Child. 2015;100(7):608–9.

37. UK Biobank. Reporting small numbers in results in research outputs using UK Biobank data. https://www.ukbiobank.ac.uk/media/ydilu0ug/numbers-for-publications-uncommon-and-geographical-variables.pdf; 2023.

38. Herdman M, Gudex C, Lloyd A, Janssen M, Kind P, Parkin D, et al. Development and preliminary testing of the new five-level version of EQ-5D (EQ-5D-5L). Qual Life Res. 2011;20(10):1727–36.

39. Ahmed N, Barnett P, Greenburgh A, Pemovska T, Stefanidou T, Lyons N, et al. Mental health in Europe during the COVID-19 pandemic: a systematic review. The Lancet Psychiatry. 2023;10(7):537–56.

40. Patel V, Fancourt D, Furukawa TA, Kola L. Reimagining the journey to recovery: The COVID-19 pandemic and global mental health. PLOS Medicine. 2023;20(4):e1004224.

41. Boorse C. The Meaning of Clinical Normality. Medical Research Archives. 2022;10(7).

42. Stein DJ, Palk AC, Kendler KS. What is a mental disorder? An exemplar-focused approach. Psychological Medicine. 2021;51(6):894–901.

43. Bishop DV. Fallibility in science: Responding to errors in the work of oneself and others. Advances in Methods and Practices in Psychological Science. 2018;1(3):432–8.

44. Kroenke K, Spitzer RL, Williams JB, Löwe B. The patient health questionnaire somatic, anxiety, and depressive symptom scales: a systematic review. General Hospital Psychiatry. 2010;32(4):345–59.

45. Kessler RC, Andrews G, Mroczek D, Ustun B, Wittchen HU. The World Health Organization composite international diagnostic interview short-form (CIDI-SF). Int J Methods Psychiatr Res. 1998;7(4):171–85.

46. Levinson D, Potash J, Mostafavi S, Battle A, Zhu X, Weissman M. Brief assessment of major depression for genetic studies: validation of CIDI-SF screening with SCID interviews. European Neuropsychopharmacology. 2017;27:S448.

47. Smith DJ, Nicholl BI, Cullen B, Martin D, Ul-Haq Z, Evans J, et al. Prevalence and characteristics of probable major depression and bipolar disorder within UK biobank: cross-sectional study of 172,751 participants. PLoS One. 2013;8(11):e75362.

48. Glaesmer H, Schulz A, Häuser W, Freyberger HJ, Brähler E, Grabe H-J. [The childhood trauma screener (CTS)-development and validation of cut-off-scores for classificatory diagnostics]. Psychiatrische Praxis. 2013;40(4):220–6.

49. Office for National Statistics. Crime in England and Wales: year ending September 2019. https://www.ons.gov.uk/peoplepopulationandcommunity/crimeandjustice/bulletins/crimeinenglandandwales/yearendingseptember2019; 2019.

50. Babor TF, Higgins-Biddle JC, Saunders JB, Monteiro MG, World Health Organization. AUDIT: The alcohol use disorders identification test: Guidelines for use in primary health care. 2001.

51. Demakakos P, Nunn S, Nazroo J. 10. Loneliness, relative deprivation and life satisfaction. In: English Longitudinal Study of Ageing, editor. Retirement, health and relationships of the older population in England (ELSA Wave 2). 297: The Institute for Fiscal Studies; 2006.

52. Smith BW, Dalen J, Wiggins K, Tooley E, Christopher P, Bernard J. The brief resilience scale: assessing the ability to bounce back. International journal of behavioral medicine. 2008;15(3):194–200.

