## Supplementary Materials (separate file) for "The UK Biobank Mental Health Enhancement 2022: Methods and Results"

Supplementary Tables 1 – 6

Supplementary Figure 1

*ST1 Characteristics of people meeting criteria for mental disorder **Female only**:*

| Characteristic | Overall | No lifetime criteria n (% have no criteria) | Depression n (% have depression) | Panic Disorder n (% have panic dx) | Any eating disorder n (% have eating dx) | Bipolar type I n (% have BP1 dx) |
|---|---|---|---|---|---|---|
| n | 97,646 | 71,372 (73%) | 22,169 (23%) | 4,913 (5.0%) | 4,447 (4.6%) | 473 (0.5%) |
| Personal characteristics | | | | | | |
| **Age**[2] | | | | | | |
| <65 | 27,915 | 17,680 (63%) | 8,475 (30%) | 2,201 (7.9%) | 2,170 (7.8%) | 237 (0.8%) |
| 65-74 | 41,461 | 30,455 (73%) | 9,307 (22%) | 1,961 (4.7%) | 1,797 (4.3%) | 179 (0.4%) |
| >75 | 28,270 | 23,237 (82%) | 4,387 (16%) | 751 (2.7%) | 480 (1.7%) | 57 (0.2%) |
| Median | 70 | 71 | 67 | 66 | 65 | 64 |
| **Sex** | | | | | | |
| Female | 97,646 | 71,372 (73%) | 22,169 (23%) | 4,913 (5.0%) | 4,447 (4.6%) | 473 (0.5%) |
| Male | 0 | 0 | 0 | 0 | 0 | 0 |
| **Ethnicity** | | | | | | |
| White | 94,576 | 69,084 (73%) | 21,510 (23%) | 4,762 (5.0%) | 4,296 (4.5%) | 450 (0.5%) |
| Black | 685 | 521 (76%) | 141 (21%) | NA[1] | NA[1] | NA[1] |
| Asian | 660 | 511 (77%) | 122 (18%) | NA[1] | NA[1] | NA[1] |
| Chinese | 261 | 218 (84%) | 36 (14%) | NA[1] | NA[1] | NA[1] |
| Mixed | 569 | 376 (66%) | 163 (29%) | NA[1] | NA[1] | NA[1] |
| Other | 543 | 414 (76%) | 110 (20%) | 151 | 151 | 23 |
| Missing | 352 | 248 (70%) | 87 (25%) | NA[1] | NA[1] | NA[1] |
| [Combined][1] | NA | NA | NA | 151 (4.9%) | 151 (4.9%) | 23 (0.1%) |
| Social factors measured at baseline | | | | | | |
| Relative deprivation[3] | 11,740 | 7,966 (68%) | 3,199 (27%) | 837 (7.1%) | 766 (6.5%) | 110 (0.9%) |
| Degree educated | 41,807 | 29,803 (71%) | 9,962 (24%) | 2,054 (4.9%) | 2,356 (5.6%) | 216 (0.5%) |
| Rents home | 4,540 | 2,759 (61%) | 1,565 (34%) | 448 (9.9%) | 344 (7.6%) | 73 (1.6%) |
| Longstanding illness | 23,652 | 15,381 (65%) | 7,272 (31%) | 1,733 (7.3%) | 1,239 (5.2%) | 255 (1.1%) |
| Smoker | 5,988 | 3,669 (61%) | 1,989 (33%) | 509 (8.5%) | 425 (7.1%) | 69 (1.2%) |
| Physical activity at least three times a week | 34,611 | 25,070 (72%) | 8,171 (24%) | 1,807 (5.2%) | 1,416 (4.1%) | 172 (0.5%) |
| Neuroticism score, mean (sd)[4] | 4.26 (3.16) | 3.71 (2.92) | 5.82 (3.29) | 6.65 (3.28) | 5.82 (3.32) | 7.34 (3.37) |
| Social factors measured in MHQ2 | | | | | | |
| Childhood adverse experience[5] | 41,575 | 26,404 (64%) | 12,871 (31%) | 3,162 (7.6%) | 2,812 (6.8%) | 360 (0.9%) |
| Adult adverse experience (lifetime) | 27,742 | 16,214 (58%) | 9,839 (35%) | 2,480 (8.9%) | 2,294 (8.3%) | 292 (1.1%) |
| Social isolation | 10,439 | 6,650 (64%) | 3,314 (32%) | 798 (7.6%) | 638 (6.1%) | 75 (0.7%) |
| Low resilience [6] | 17,838 | 8,223 (46%) | 8,586 (48%) | 2,314 (13%) | 1,497 (8.4%) | 270 (1.5%) |
| Loneliness score, median (IQR) [7] | 3 (3-5) | 3 (3-4) | 4 (3-6) | 5 (3-6) | 4 (3-6) | 5 (3-7) |
| Self-rated health median (IQR) [8] | 83 (72-91) | 85 (75-92) | 80 (64-89) | 75 (60-86) | 80 (66-90) | 69 (45-82) |

(1) To preserve participant privacy, non-White and missing ethnicities are collapsed to a single value ("Combined") if one cell size is smaller than 10.
(2) Age at MHQ2 is at date of completion of MHQ2
(3) Based on Townsend Deprivation Score of residence, where deprived ≥+2 (0 is average, and higher scores are more deprived)
(4) Any domain positive on the Childhood Trauma Screener
(5) Brief Resilience Scale, low = 1 to 2.99, where scale is 1-5
(6) Scores on at least two of three isolation questions
(7) Loneliness score from 3 to 9 points, with 3 being least lonely
(8) EQ-5D visual analogue scale, from 0 to 100, higher indicating better health

*ST2 Characteristics of people meeting criteria for mental disorder **Male only***

| Characteristic | Overall | No lifetime criteria n (% have no criteria) | Depression n (% have depression) | Panic Disorder n (% have panic dx) | Any eating disorder n (% have eating dx) | Bipolar type I n (% have BP1 dx) |
|---|---|---|---|---|---|---|
| n | 71,607 | 61,603 (86%) | 9,074 (13%) | 1,790 (2.5%) | 315 (0.4%) | 248 (0.3%) |
| Personal characteristics | | | | | | |
| **Age**[2] | | | | | | |
| <65 | 17,981 | 14,142 (79%) | 3,457 (19%) | 812 (4.5%) | 167 (0.9%) | 115 (0.6%) |
| 65-74 | 29,001 | 24,988 (86%) | 3,655 (13%) | 681 (2.3%) | 102 (0.4%) | 85 (0.3%) |
| >75 | 24,625 | 22,473 (91%) | 1,962 (8.0%) | 297 (1.2%) | 46 (0.2%) | 48 (0.2%) |
| Median | 71 | 72 | 67 | 66 | 64 | 66 |
| **Sex** | | | | | | |
| Female | 0 | 0 | 0 | 0 | 0 | 0 |
| Male | 71,607 | 61,603 (86%) | 9,074 (13%) | 1,790 (2.5%) | 315 (0.4%) | 248 (0.3%) |
| **Ethnicity** | | | | | | |
| White | 69,219 | 59,521 (86%) | 8,796 (13%) | 1,731 (2.5%) | 302 (0.4%) | 232 (0.3%) |
| Black | 438 | 400 (91%) | 35 (8.0%) | NA[1] | NA[1] | NA[1] |
| Asian | 788 | 721 (91%) | 62 (7.9%) | NA[1] | NA[1] | NA[1] |
| Chinese | 114 | 101 (89%) | 13 (11%) | NA[1] | NA[1] | NA[1] |
| Mixed | 294 | 227 (77%) | 60 (20%) | NA[1] | NA[1] | NA[1] |
| Other | 359 | 299 (83%) | 54 (15%) | NA[1] | NA[1] | NA[1] |
| Missing | 395 | 334 (85%) | 54 (14%) | NA[1] | NA[1] | NA[1] |
| [Combined][1] | NA | NA | NA | 59 (2.5%) | 13 (0.5%) | 16 (0.7%) |
| Social factors measured at baseline | | | | | | |
| Relative deprivation[3] | 8,275 | 6,693 (81%) | 1,430 (17%) | 319 (3.9%) | 82 (1.0%) | 55 (0.7%) |
| Degree educated | 33,504 | 28,762 (86%) | 4,318 (13%) | 764 (2.3%) | 166 (0.5%) | 124 (0.4%) |
| Rents home | 3,369 | 2,490 (74%) | 800 (24%) | 193 (5.7%) | 50 (1.5%) | 37 (1.1%) |
| Longstanding illness | 20,644 | 16,720 (81%) | 3,575 (17%) | 780 (3.8%) | 153 (0.7%) | 139 (0.7%) |
| Smoker | 5,873 | 4,727 (80%) | 1,035 (18%) | 242 (4.1%) | 39 (0.7%) | 47 (0.8%) |
| Physical activity at least three times a week | 27,760 | 23,829 (86%) | 3,570 (13%) | 706 (2.5%) | 122 (0.4%) | 91 (0.3%) |
| Neuroticism score, mean (sd)[4] | 3.36 (3.10) | 3.01 (2.89) | 5.55 (3.44) | 6.49 (3.56) | 6.46 (3.65) | 6.46 (3.45) |
| Social factors measured in MHQ2 | | | | | | |
| Childhood adverse experience[5] | 29,474 | 23,766 (81%) | 5,196 (18%) | 1,118 (3.8%) | 207 (0.7%) | 176 (0.6%) |
| Adult adverse experience (lifetime) | 10,285 | 7,370 (72%) | 2,626 (26%) | 644 (6.3%) | 131 (1.3%) | 107 (1.0%) |
| Social isolation | 6,926 | 5,449 (79%) | 1,361 (20%) | 279 (4.0%) | 68 (1.0%) | 44 (0.6%) |
| Low resilience [6] | 8,529 | 4,880 (57%) | 3,358 (39%) | 813 (9.5%) | 143 (1.7%) | 136 (1.6%) |
| Loneliness score, median (IQR) [7] | 3 (3-5) | 3 (3-4) | 4 (3-6) | 5 (3-6) | 5 (3-7) | 5 (3-7) |

| | | | | | | |
|---|---|---|---|---|---|---|
| Self-rated health median (IQR) [8] | 84 (74-90) | 85 (75-90) | 79 (65-88) | 74 (55-85) | 73 (50-85) | 71 (51-85) |

(1) To preserve participant privacy, non-White and missing ethnicities are collapsed to a single value ("Other") if one cell size is smaller than 10.
(2) Age at MHQ2 is at date of completion of MHQ2
(3) Based on Townsend Deprivation Score of residence, where deprived ≥ +2 (where 0 is average, and higher scores are more deprived)
(4) Any domain positive on the Childhood Trauma Screener
(5) Brief Resilience Scale, low = 1 to 2.99, where scale is 1-5
(6) Scores on at least two of three isolation questions
(7) Loneliness score from 3 to 9 points, with 3 being least lonely
(8) EQ-5D visual analogue scale, from 0 to 100, higher indicating better health

*ST3 Age*- and sex-stratified results for cases of depression, anxiety and harmful alcohol use in those who completed both questionnaires according to measures of current mental health (PHQ-9 and GAD-7 "last two weeks", AUDIT "in the last year")*

| **Instrument** | **Depression (PHQ-9) case** | | | | **Anxiety (GAD-7) case** | | | | **Harmful alcohol use (AUDIT) case** | | | |
|---|---|---|---|---|---|---|---|---|---|---|---|---|
| **Sex** | **M** | | **F** | | **M** | | **F** | | **M** | | **F** | |
| **Year** | **2016** | **2022** | **2016** | **2022** | **2016** | **2022** | **2016** | **2022** | **2016** | **2022** | **2016** | **2022** |
| **Overall N** | 46,800 | 46,800 | 64,475 | 64,475 | 46,800 | 46,800 | 64,475 | 64,475 | 46,800 | 46,800 | 64,475 | 64,475 |
| Positive | 957 | 995 | 1670 | 1809 | 1400 | 1318 | 3118 | 3106 | 1891 | 1588 | 1155 | 968 |
| Positive % | 2.0% | 2.1% | 2.6% | 2.8% | 3.0% | 2.8% | 4.8% | 4.8% | 4.0% | 3.4% | 1.8% | 1.5% |
| **Under age 52 N** | 3,132 | 0 | 4,820 | 0 | 3,132 | 0 | 4,820 | 0 | 3,132 | 0 | 4,820 | 0 |
| Positive | 125 | 0 | 236 | 0 | 188 | 0 | 357 | 0 | 215 | 0 | 153 | 0 |
| Positive % | 4.0% | NA | 4.9% | NA | 6.0% | NA | 7.4% | NA | 6.9% | NA | 3.2% | NA |
| **Age group 52-58 N** | 8,358 | 4,048 | 13,660 | 6,184 | 8,358 | 4,048 | 13,660 | 6,184 | 8,358 | 4,048 | 13,660 | 6,184 |
| Positive | 299 | 186 | 579 | 346 | 407 | 256 | 912 | 529 | 482 | 252 | 369 | 203 |
| Positive % | 3.6% | 4.6% | 4.2% | 5.6% | 4.9% | 6.3% | 6.7% | 8.6% | 5.8% | 6.2% | 2.7% | 3.3% |
| **Age group 59-65 N** | 12,130 | 8,728 | 18,631 | 14,348 | 12,130 | 8,728 | 18,631 | 14,348 | 12,130 | 8,728 | 18,631 | 14,348 |
| Positive | 285 | 282 | 487 | 551 | 383 | 354 | 902 | 870 | 544 | 444 | 381 | 335 |
| Positive % | 2.3% | 3.2% | 2.6% | 3.8% | 3.2% | 4.1% | 4.8% | 6.1% | 4.5% | 5.1% | 2.0% | 2.3% |
| **Age group 66-72 N** | 16,459 | 12,614 | 20,450 | 19,034 | 16,459 | 12,614 | 20,450 | 19,034 | 16,459 | 12,614 | 20,450 | 19,034 |
| Positive | 177 | 240 | 306 | 450 | 322 | 325 | 764 | 817 | 520 | 473 | 220 | 278 |
| Positive % | 1.1% | 1.9% | 1.5% | 2.4% | 2.0% | 2.6% | 3.7% | 4.3% | 3.2% | 3.7% | 1.1% | 1.5% |
| **Age group 73-79 N** | 6,704 | 16,332 | 6,904 | 19,695 | 6,704 | 16,332 | 6,904 | 19,695 | 6,704 | 16,332 | 6,904 | 19,695 |
| Positive | 71 | 225 | 62 | 362 | 100 | 299 | 183 | 727 | 130 | 351 | 32 | 135 |
| Positive % | 1.1% | 1.4% | 0.9% | 1.8% | 1.5% | 1.8% | 2.7% | 3.7% | 1.9% | 2.1% | 0.5% | 0.7% |
| **Age group >80 N** | 17 | 5,076 | 10 | 5,207 | 17 | 5,076 | 10 | 5,207 | 17 | 5,076 | 10 | 5,207 |
| Positive | 0 | 62 | 0 | 100 | 0 | 84 | 0 | 163 | 0 | 68 | 0 | 17 |
| Positive % | NA | 1.2% | NA | 1.9% | NA | 1.7% | NA | 3.1% | NA | 1.3% | NA | 0.3% |

* Age at MHQ2 is at date of completion (NA where this was missing, n=28), age at MHQ1 is at approximate date of completing MHQ1

*ST4: Relative risks of being a case on current mental health outcomes in 2022 compared to 2016 in age* and sex specific groups in 2022 in people who completed both MHQs. See ST3 for sample size by age and sex.*

| | PHQ-9 Depression | | GAD-7 Anxiety | | Harmful alcohol use | |
|---|---|---|---|---|---|---|
| | M (n=46,800) | F (n=64,475) | M (n=46,800) | F (n=64,475) | M (n=46,800) | F (n=64,475) |
| Age* and sex specific **relative risk** positive result | | | | | | |
| <52 | NA | NA | NA | NA | NA | NA |
| 52-58 | 1.28 | 1.33 | 1.29 | 1.28 | 1.07 | 1.22 |
| 59-65 | 1.39 | 1.46 | 1.28 | 1.27 | 1.13 | 1.15 |
| 66-72 | 1.73 | 1.60 | 1.30 | 1.16 | 1.16 | 1.36 |
| 73-79 | 1.27 | 2.00 | 1.20 | 1.37 | 1.11 | 1.40 |
| >79 | NA | NA | NA | NA | NA | NA |
| Sex specific **relative risk** positive result (including all age groups) | | | | | | |
| All ages | 1.05 | 1.08 | 0.93 | 1.00 | 0.85 | 0.83 |
| Overall | | | | | | |
| All ages and sex | 1.07 | | 0.98 | | 0.84 | |

* Age at MHQ2 is at date of completion (NA where this was missing, n=28), age at MHQ1 is at approximate date of completing MHQ1

*ST5 and ST6 Comparison of lifetime conditions by participants, based on meeting criteria in MHQ1 and MHQ2 (restricted to those participants who answered both questionnaires)*

*ST5 Comparison of lifetime conditions by participants, based on meeting criteria in MHQ1 and MHQ2,* ***Female only***

| Case criteria | Met in both | Met 1 not 2 | Met 2 not 1 * | Neither | Agree (%) | Kappa | New onset |
|---|---|---|---|---|---|---|---|
| Depression | 10603 | 7850 | 4039 | 41983 | 82% | 52% | 1001 (25%) |
| Bipolar | 102 | 255 | 176 | 63942 | 99% | 32% | 19 (11%) |
| Self-harm | 2540 | 938 | 1266 | 59731 | 97% | 68% | 275 (22%) |
| Cannabis | 11124 | 1995 | 1962 | 49394 | 94% | 81% | 162 (8.3%) |
| Any SR clinician diagnosis | 17879 | 6877 | 2802 | 36917 | 85% | 67% | NA |

*Met 1 = met criteria for lifetime phenotype in MHQ1, Met 2 = met criteria for lifetime phenotype in MHQ2, New = of those who met criteria in 2 and not 1, how many reported that their first experience was after the MHQ1 questionnaire or onset was unknown (was not asked with regards to clinician diagnosis)*

*ST6 Comparison of lifetime conditions by participants, based on meeting criteria in MHQ1 and MHQ2,* ***Male only***

| Case criteria | Met in both | Met 1 not 2 | Met 2 not 1 * | Neither | Agree (%) | Kappa | New onset |
|---|---|---|---|---|---|---|---|
| Depression | 4009 | 3820 | 1908 | 37063 | 88% | 51% | 501 (26%) |
| Bipolar | 55 | 193 | 103 | 46449 | 99% | 27% | 10 (10%) |
| Self-harm | 958 | 473 | 522 | 44847 | 98% | 65% | 135 (26%) |
| Cannabis | 10506 | 2020 | 1431 | 32843 | 93% | 81% | 92 (6.4%) |
| Any SR clinician diagnosis | 7822 | 4801 | 1484 | 32693 | 87% | 63% | NA |

*Met 1 = met criteria for lifetime phenotype in MHQ1, Met 2 = met criteria for lifetime phenotype in MHQ2, New = of those who met criteria in 2 and not 1, how many reported that their first experience was after the MHQ1 questionnaire or onset was unknown (was not asked with regards to clinician diagnosis)*

Supplementary Figure 1

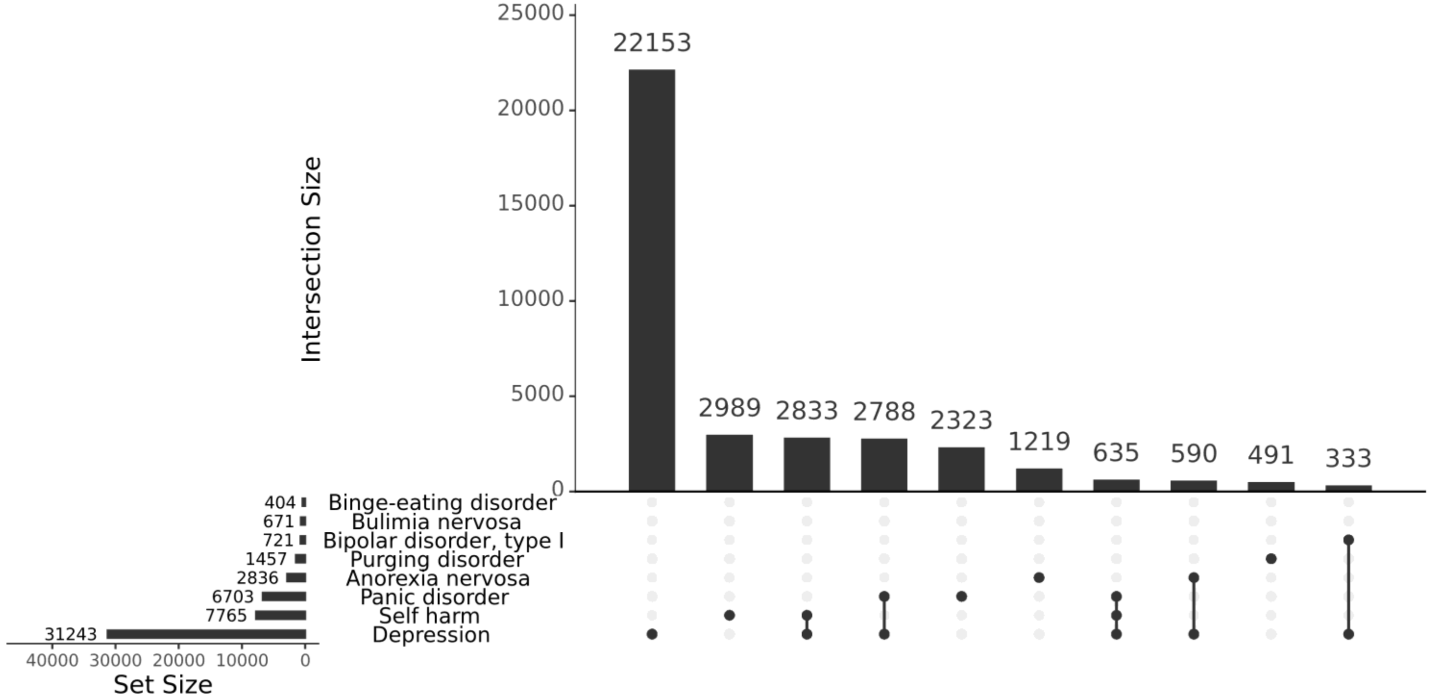


*UPSET plots show the relative proportion and degree of intersection between concepts, in this case, phenotypes defined by symptom-based lifetime criteria in the MHQ2: depression, bipolar affective disorder type 1, panic disorder, and the four eating disorders, plus self-reported self-harm ever. The ten most common intersections are shown (out of a potential 127), which are mutually exclusive; for example, if a respondent met the criteria for lifetime depression, bipolar disorder and self-harm, they will not appear in the depression and self-harm or the depression and bipolar disorder intersects.*

Supplementary Figure 1: An UPSET plot of lifetime phenotypes in MHQ2 showing the ten most common 'combinations'.
